# The Equilibrium and Pandemic Waves of COVID-19 in the US

**DOI:** 10.1101/2023.02.13.23285847

**Authors:** Zixin Hu, Xiaoxi Hu, Tao Xu, Kai Zhang, Henry H Lu, Jinying Zhao, Eric Boerwinkle, Li Jin, Momiao Xiong

## Abstract

**Importance:** Removing the epidemic waves and reducing the instability level of an endemic critical point of COVID-19 dynamics are fundamental to the control of COVID-19 in the US.

**Objective:** To develop new mathematic models and investigate when and how will the COVID-19 in the US be evolved to endemic.

**Design, Setting, and Participants:** To solve the problem of whether mass vaccination against SARS-CoV-2 will ultimately end the COVID-19 pandemic, we defined a set of nonlinear ordinary differential equations as a mathematical model of transmission dynamics of COVID-19 with vaccination. Multi-stability analysis was conducted on the data for the daily reported new cases of infection from January 12, 2021 to December 12, 2022 across 50 states in the US using the developed dynamic model of COVID-19 and limit cycle theory.

**Main Outcomes and Measures:** Eigenvalues and the reproduction number under the disease-free equilibrium point and endemic equilibrium point were used to assess the stability of the disease-free equilibrium point and endemic equilibrium point. Both analytic analysis and numerical methods were used to determine the instability level of new cases of COVID-19 in the US under the different types of equilibrium points and to investigate how the system moves back and forth between stable and unstable states of the system and how the pandemic COVD-19 will evolve to endemic in the US.

**Results:** Multi-stability analysis identified two types of critical equilibrium points, disease-free endemic equilibrium points in the COVID-19 transmission dynamic system. The transmissional, recovery, vaccination rates and vaccination effectiveness during the major transmission waves of COVID-19 across 50 states in the US were estimated. These parameters in the model varied over time and across the 50 states. The eigenvalues and the reproduction numbers *R*_0_ and 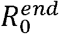 in the disease-free equilibrium point and endemic equilibrium point were estimated to assess stability and classify equilibrium points. They also varied from state to state. The impacts of the transmission and vaccination parameters on the stability of COVID-19 were simulated, and stability attractor regions of these parameters were found and ranked for all 50 states in the US. The US experienced five major epidemic waves, endemic equilibrium points of which across 50 states were all in unstable states. However, the combination of re-infection and vaccination (hybrid immunity) may provide strong protection against COVID-19 infection, and stability analysis showed that these unstable equilibrium points were toward stable points. Theoretical analysis and real data analysis showed that additional epidemic waves may be possible in the future, but COVID-19 across all 50 sates in the US is rapidly moving toward stable endemicity.

**Conclusions and Relevance:** Both stability analysis and observed epidemic waves in the US indicated that the pandemic might not end with the disappearance of the virus. However, after enough people gained immune protection from vaccination and from natural infection, COVID-19 would become an endemic disease, as the stability analysis showed. Educating the population about multiple epidemic waves of the transmission dynamics of COVID-19 and designing optimal vaccine rollout are crucial for controlling the pandemic of COVID-19 and its evolving to endemic.

**Key Points:** *Questio:* The US has already experienced five waves of the epidemic. We urgently need to know when and how will COVID-19 be evolved into endemic.

*Findings:* To solve the problem, we developed a mathematical model of transmission dynamics of COVID-19 with vaccination and performed a multi-stability analysis of COVID-19 transmission dynamics in the US. We found that COVID-19 dynamics of all 50 states in the US were getting closer and closer to endemic and stable states.

*Meaning:* COVID-19 dynamics of all 50 states in the US are toward stable states and will be evolved to endemic in the near future.

## 1. Introduction

Since the outbreaks of COVID-19 in December 2019 in Wuhan, China, the US has experienced five waves over the summer of 2020, winter months of 2020-2021, and summer-fall months of 2021, winter months of 2021-2022 and summer months of 2022. As of January 4, 2023, a total of 101,094,670 COVID-19 cases have been observed in the US. The current 7-day (as of January 4, 2023) average of weekly new cases (67,243) increased by 16.2% compared with the previous 7-day average (57,847).^1^ Will the US experience a sixth wave? What causes the multiple waves in the US and can we end the pandemic with massive vaccinations and reinfections? Many factors contribute to the multiple waves, such as the immune escape of the emergent virus variants and natural immune escape of infection, lifted or poorly adhered non-pharmaceutical interventions (NPI), vaccination and the population effectiveness of vaccines over time, and population socio-economic, age, and geographic structures.^2^ An important factor that is often ignored is the inherent instability behavior of the transmission dynamic system of COVID-19. Understanding the causation factors of such multiple waves and designing the appropriate strategies to end the pandemic are crucial to our control of the spread of COVID-19.

There is hope for ending the spread of COVID-19 and returning to normal through vaccination and natural immunity, and appropriate and flexible NPI. Unfortunately, mutations and natural selection generate new variants that increase virus replication, transmission, and escape of the immune system. These new variants raise concerns for increased transmission and escape from both vaccine and natural infection immunity.^3^

The COVID-19 pandemic is devastating. The total number of cases and deaths in the world reached 660,131,952 and 6,690,473, respectively, by January 10, 2023.^4^ Instead of global eradication of COVID-19 very soon, we observed its multiple epidemic waves. Developing a mechanic analytic model for COVID-19 transmission dynamics is essential for periodically investigating unstable patterns of COVID-19 dynamics, uncovering the major factors underlying the multiple epidemic waves, and better understanding the challenges we are facing in eliminating COVID-19. Widely used mathematical models for the transmission dynamics of COVID-19 are epidemic compartment models.^5-11^ Although these models cannot accurately predict the transmission dynamics due to the imprecise description of biological processes and limited data resources, they still can roughly capture the dynamic patterns and features of COVID-19. These mathematical models are often described by a set of differential equations, however, only estimating the parameters in the equations from the real data by numerically solving these differential equations are not good enough to capture the dynamic features of the COVID-19 transmissions and design strategies to eliminate COVID-19. Stability analysis, which is often overlooked in epidemic models of COVID-19, is essential to explore dynamic systems of COVID-19 transmissions. COVID-19 periodically changes between stationary and nonstationary states, which results in multiple epidemic waves.

To address these issues, we will extend the epidemic SEIR model to incorporate an additional vaccination compartment. In other words, we will consider five stages of infection: Susceptible (S), exposed (E), infection (I), recovered (R), and vaccinated (V). The new model is denoted as SEIRV. We will focus on multiple stability analysis of SEIRV for COVID-19.^10-14^ We will identify its disease-free and epidemic critical equilibrium points and further use the next-generation matrix methods to calculate the reproduction numbers for the disease-free critical point (*R*_0_) and the epidemic critical point 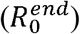. Armed with *R*_0_ and 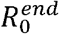, we will use stability theory to derive the conditions leading to stable states or unstable states of the SEIRV dynamic system. We will further identify the attractors that determine the range of the parameters underlying the stable states of the covid-19 dynamic system. The limit cycle^15^ of the COVID-19 dynamic system will be investigated and the mechanism underlying the epidemic waves will be uncovered. Finally, based on the multiple stability analysis, we will provide information for designing strategies to mitigate and finally end the spread of COVID-19 in the US.

## 2. Methods

### 2.1. Data sources

The number of cases and deaths for each state from January 21, 2020 to December 12, 2022 was downloaded from https://github.com/nytimes/covid-19-data. The vaccination data, including the number of vaccines distributed, the number of people who received at least one shot of vaccinations and full vaccination for each state from January 12, 2021 to December 12, 2022 were downloaded from https://ourworldindata.org/us-states-vaccinations.

### 2.2. SEIRV compartment model for COVID-19 transmission dynamics

We constructed a compartmental model based on a deterministic system of nonlinear differential equations for COVID-19 transmission dynamics, which further takes into account vaccination compared with previous publications. The architecture of the model is shown in Figure 1. It includes the susceptible (*S*(*t*)), exposed (*E*(*t*)), infected (*I*(*t*)), recovered (*R*(*t*)), and vaccinated (*V*(*t*)) individuals, each one forming a compartment. The model assumes that the transition of individuals from one compartment to another depends on the stage of the disease. The model also assumes a constant recruiting rate (births) Λ to the susceptible individuals (*S*(*t*)), natural death rate *μ*, transmission rate *β*, vaccination rate *α*, incubation rate *γ*, the probability of the recovery or death *δ*, and vaccine inefficiency *σ* (0 ≤ *σ* ≤ 1) where 1 − *σ* represents the population vaccine efficacy. Five nonlinear differential equations defined in equations (S1-S5) are used to model the transmission dynamics of *S*(*t*), *E*(*t*), *I*(*t*), *R*(*t*) and *V*(*t*). The differential equations start with non-negative initial conditions *S*(0), *E*(0), *I*(0), *R*(0) and *V*(0). We assume that *N* = *S*(*t*) + *E*(*t*) + *I*(*t*) + *R*(*t*) + *V*(*t*) is the total population size.

**Figure 1.**
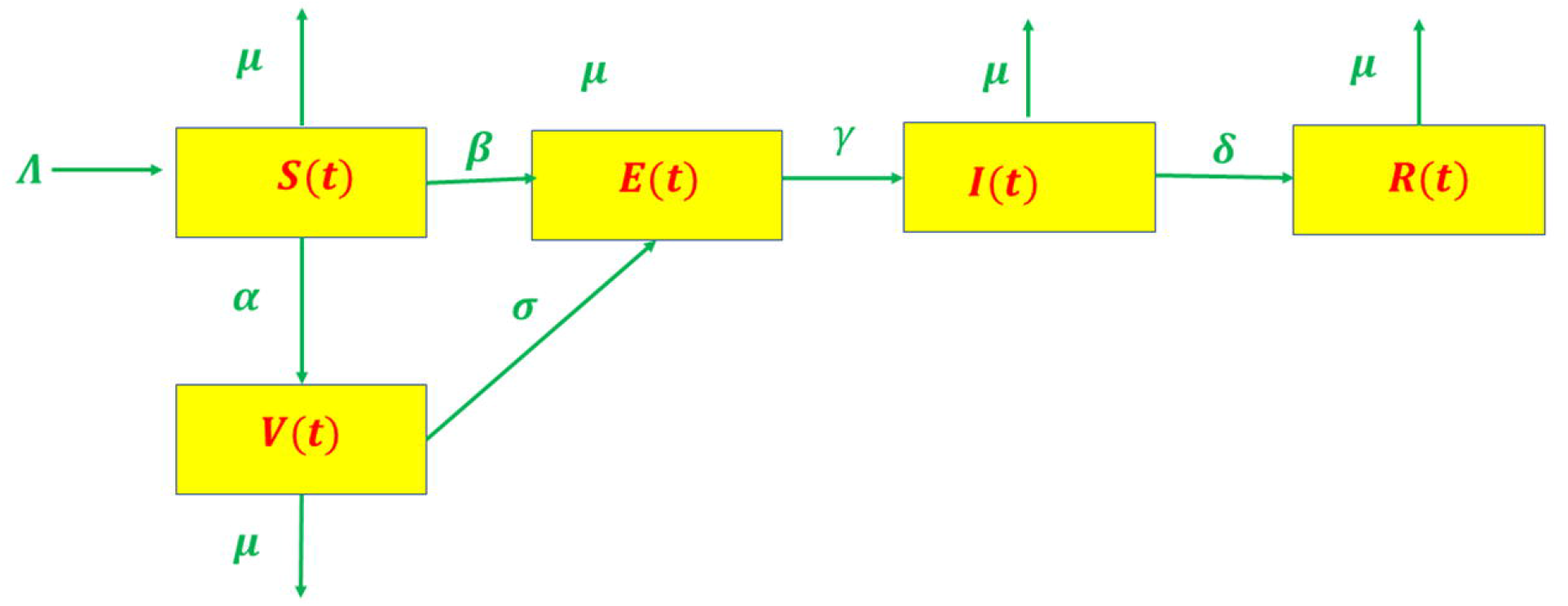
Dynamic transmission model of COVID-19.

### 2.3. The properties of solutions

We show in Supplementary 2 that all variables *S*(*t*), *E*(*t*), *I*(*t*), *R*(*t*), V(*t*) are bounded in the region Ω:

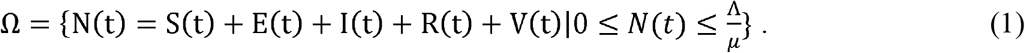

### 2.4. Reproduction number

The basic reproduction number, denoted as *R*_0_, is an important threshold quantity which determines whether the COVID-19 will continuously spread in the population or disappear. It is defined as the average number of secondary cases produced by one infected individual introduced into a population of susceptible individuals.^16^ We compute the basic reproduction number *R*_0_ using the next generation matrix method as (Supplementary 3.1)

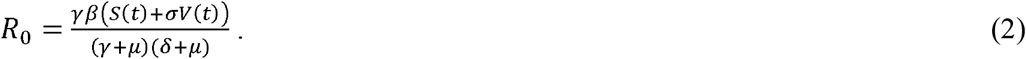

The reproduction number *R*_0_ is used to measure the transmission potential of a disease. Intuitively, we can expect that if *R*_0_ < 1 then the number of new cases of COVID-19 will decrease, and the number of new cases will increase if *R*_0_ > 1.

### 2.5. Steady sate analysis of SEIRV for COVID-19 transmission dynamic system

Predictions of multistable structural dynamics are paramount to the development and implementation of vaccination and population public health intervention for controlling the spread of COVID-19 under highly uncertain biological, economic, political, and environmental perturbation. Although a direct numerical solution to differential equations can be used to calculate the steady state, analytic analysis of the steady state can reveal how the parameters affect the steady state and enable prediction of near- and far-from equilibrium response. Such analysis will provide useful information on the endemic waves of COVID-19 and explore the strategies for mitigating or ending COVID-19.

COVID-19 transmission dynamic system is an autonomous system in which the independent time variable *t* does not explicitly appear in the differential equations. We will focus on steady state and stability analysis of autonomous systems to investigate the qualitative dynamic behavior of COVID-19 dynamic systems. The central issue in stability analysis is to identify isolated critical (equilibrium) points of COVID-19 transmission dynamics. The isolated critical points can be classified as disease-free (number of new cases is zero) critical point and endemic (new cases are not zero) critical point. Two classes of critical points are given as follows (For details, please see Supplementary materials 3.2).

1. Disease-free critical (equilibrium) point:

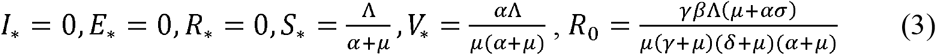
2. Endemic critical (equilibrium) point:
3. 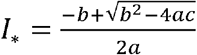,
4. 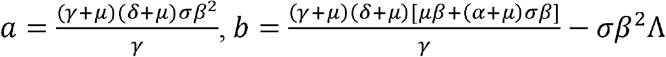,
5. 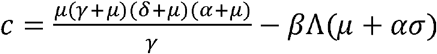,
6. 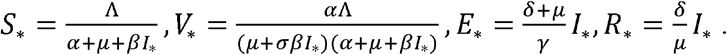.
7. 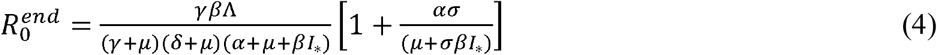

Now we study the properties of the critical points which determine whether the system is stable or unstable or whether the number of new cases decreases or increases. Disease-free and endemic critical points are separately investigated and briefly presented (For details, please see Supplementary materials 3.3).

#### Disease-free critical (equilibrium) point

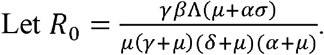

#### The disease-free critical point can be classified as three cases

1. when *R*_0_ > 1, the disease-free critical point is classified as a asymptotically stable node;
2. when *R*_0_ = 1, the disease-free critical point is classified as an unstable node; and
3. when *R*_0_ < 1, the disease-free critical point is classified as an unstable saddle point.

#### Endemic critical (equilibrium) point

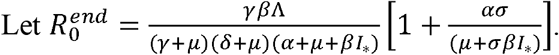

Then, in summary, the **endemic equilibrium point can be classified into three cases:**

1. when 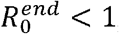, the endemic equilibrium point is classified as an asymptotically stable node;
2. when 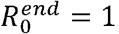, the endemic equilibrium point is classified as an unstable node; and
3. when 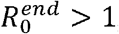, the endemic equilibrium point is classified as an unstable saddle point.

### 2.6. Parameter estimation

We can use the steady states of the COVID-19 transmission dynamic system to estimate the parameters in the nonlinear differential equation models and then use the estimated parameters to identify and classify the equilibrium points.

Let

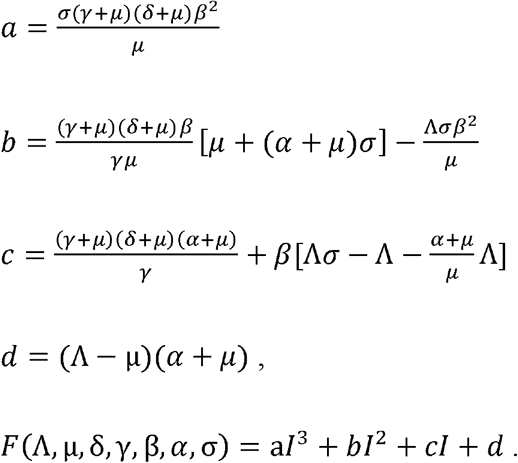

The parameters are estimated by minimizing

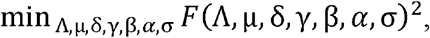

where *I* is the observed number of new cases in the steady state (hypothesized) of COVID-19 dynamic system (S1-S5).

## 3. Real data analysis

### 3.1. Estimation of parameters

We start data with January 12, 2021 and end data with December 12, 2022. To study endemic equilibrium points, we consider three steady state periods: (1) April – July, 2021; (2) March – May, 2022; and (3) September – November, 2022 (Figure S1). We used the data for the daily reported new cases of infection to fit the COVID-19 model for estimating the model parameters. The estimated median values and standard deviation of the parameters *α, β, γ, δ*, and *σ* in three steady periods of 50 states in the US were shown in Figure 2. We observed in Figure 2 that the estimated parameters *β, σ, γ*, and *δ* decreased, while parameter *α* increased from time periods April – July, 2021 to September – November, 2022. This showed that vaccination rates increased while all parameters related to the transmission of COVID-19 decreased, which implied that the spread rates of COVID-19 were gradually reduced due to increased vaccinations in the US.

**Figure 2.**
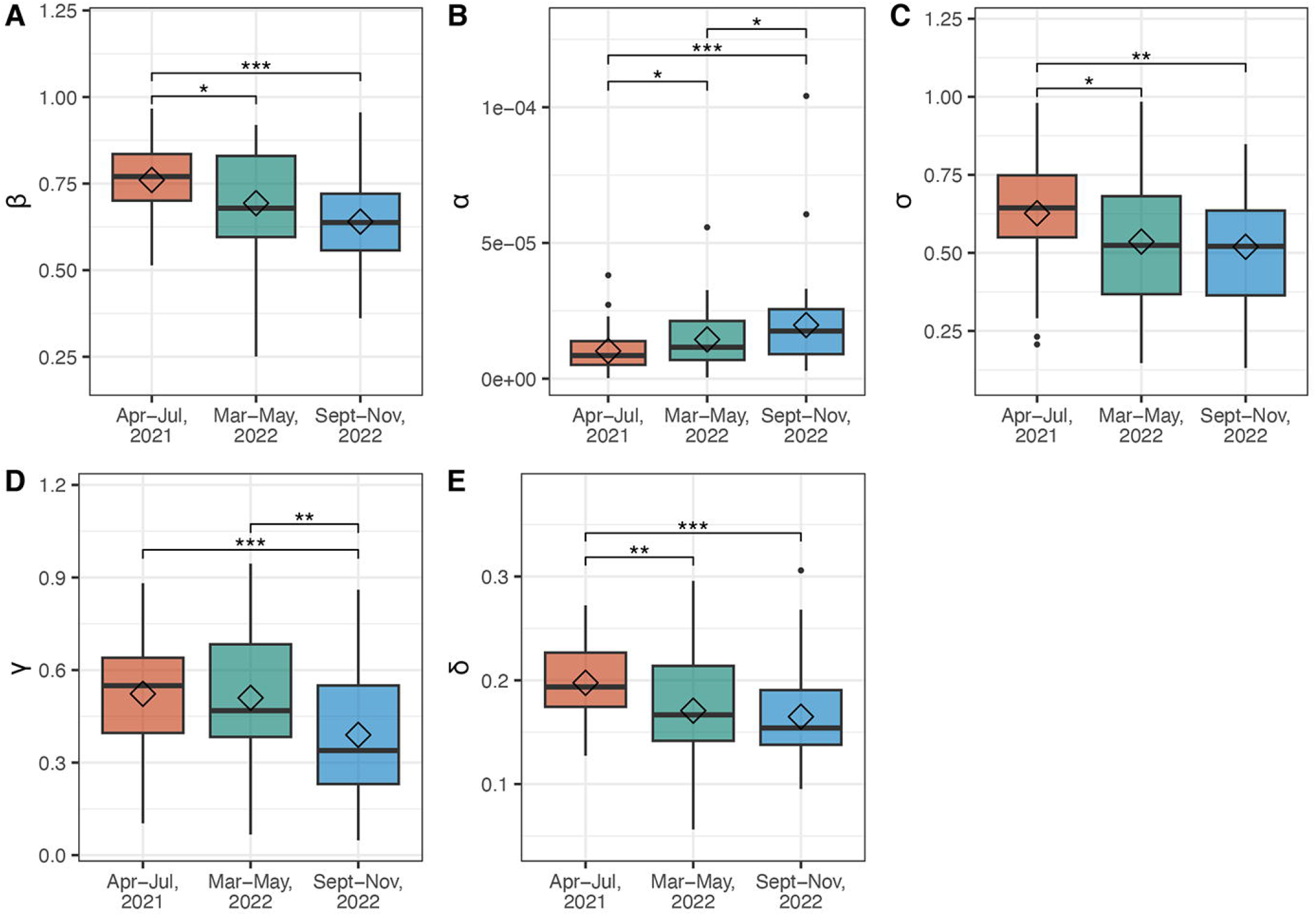
Box plot for the parameters β, *α, σ, γ, δ* in three steady periods (April – July, 2021; March – May 2022; September – November, 2022) across 50 states in the US. (A) Box plot for the parameter *β*, (B) Box plot for the parameter *α*, (C) Box plot for the parameter *σ*, (D) Box plot for the parameter *γ* and (E) Box plot for the parameter *δ*.

### 3.2. Eigenvalues, reproduction number at the endemic equilibrium points and stability analysis

To assess the stability in the steady states of the dynamics of COVID-19 across 50 states in the US, we calculated the eigenvalues of the Jacobian matrix at the endemic equilibrium points. The total number of eigenvalues in the system was five. However, theoretical analysis of stability showed that one of the eigenvalues was always negative. We observed that at least one eigenvalue was positive in all three steady periods across 50 states (Table 1). This shows that until now no dynamics of COVID-19 across 50 states in the US have reached stable states. However, the largest eigenvalue, which determined whether the COVID-19 transmission dynamic system at the endemic equilibrium point was stable or unstable, decreased in 32 states when the system went from the first steady period (April – July, 2021) to the third steady period (September – November, 2022) (Table 1), and did not show significant changes in 3 states during three steady periods.

**Table 1.** The largest eigenvalue at the endemic equilibrium points in three steady periods across 50 states in the US.

|  | April - July, 2021 | March - May, 2022 | September - November, 2022 |
| --- | --- | --- | --- |
| state | $\lambda_4$ | $\lambda_4$ | $\lambda_4$ |
| AK | 1.56E-01 | 6.10E-02 | 1.90E-01 |
| AL | 1.71E-01 | 1.17E-01 | 1.12E-01 |
| AR | 1.22E-01 | 2.61E-01 | 6.73E-02 |
| AZ | 1.35E-01 | 6.94E-02 | 1.36E-01 |
| CA | 2.11E-01 | 1.18E-01 | 1.55E-01 |
| CO | 1.36E-01 | 6.24E-02 | 9.90E-02 |
| CT | 1.32E-01 | 1.53E-01 | 1.10E-01 |
| DE | 2.41E-01 | 1.00E-01 | 9.05E-02 |
| FL | 1.29E-01 | 1.41E-01 | 1.36E-01 |
| GA | 9.10E-02 | 1.11E-01 | 1.80E-01 |
| HI | 1.85E-01 | 1.05E-01 | 1.62E-01 |
| IA | 1.72E-01 | 1.59E-01 | 1.74E-01 |
| ID | 1.19E-01 | 1.86E-01 | 1.11E-01 |
| IL | 8.04E-02 | 1.36E-01 | 1.24E-01 |
| IN | 9.64E-02 | 3.55E-01 | 1.46E-01 |
| KS | 1.66E-01 | 1.57E-01 | 1.86E-02 |
| KY | 1.83E-01 | 9.89E-02 | 8.74E-02 |
| LA | 8.99E-02 | 2.25E-01 | 1.66E-01 |
| MA | 1.90E-01 | 7.80E-02 | 7.56E-02 |
| MD | 1.78E-01 | 3.46E-01 | 1.00E-01 |
| ME | 2.96E-01 | 9.65E-02 | 1.42E-01 |
| MI | 3.05E-01 | 2.14E-01 | 5.51E-02 |
| MN | 2.36E-01 | 1.84E-01 | 9.80E-02 |
| MO | 5.65E-02 | 2.22E-01 | 1.50E-01 |
| MS | 1.35E-01 | 2.10E-01 | 1.40E-01 |
| MT | 1.33E-01 | 2.96E-01 | 9.10E-02 |
| NC | 1.10E-01 | 8.09E-02 | 6.41E-02 |
| ND | 2.02E-01 | 2.29E-01 | 7.88E-02 |
| NE | 2.20E-01 | 2.11E-01 | 2.56E-01 |
| NH | 2.73E-01 | 1.41E-01 | 1.59E-01 |
| NJ | 2.75E-01 | 1.26E-01 | 7.17E-02 |
| NM | 1.80E-01 | 2.01E-01 | 8.91E-02 |
| NV | 5.38E-02 | 1.87E-01 | 2.70E-01 |
| NY | 2.23E-01 | 1.75E-01 | 5.63E-02 |
| OH | 1.84E-01 | 2.50E-01 | 8.04E-02 |
| OK | 1.73E-01 | 1.27E-01 | 1.24E-01 |
| OR | 2.36E-01 | 1.33E-01 | 1.20E-01 |
| PA | 2.99E-01 | 3.09E-01 | 1.13E-01 |
| PR | 1.78E-01 | 1.08E-01 | 5.07E-02 |
| RI | 6.73E-02 | 8.17E-02 | 6.55E-02 |
| SC | 1.40E-01 | 2.51E-01 | 1.21E-01 |
| SD | 2.08E-01 | 2.25E-01 | 9.39E-02 |
| TN | 1.38E-01 | 2.35E-01 | 8.61E-02 |
| TX | 1.15E-01 | 2.99E-02 | 2.14E-01 |
| UT | 9.34E-02 | 2.78E-01 | 1.31E-01 |
| VA | 1.72E-01 | 8.09E-02 | 1.18E-01 |
| VT | 1.44E-01 | 6.34E-02 | 1.18E-01 |
| WA | 1.78E-01 | 6.34E-02 | 1.30E-01 |
| WI | 1.52E-01 | 1.43E-01 | 7.52E-02 |
| WV | 2.29E-01 | 2.19E-01 | 8.74E-02 |
| WY | 1.86E-02 | 2.22E-01 | 1.05E-01 |

Although the reproduction number 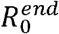 at the endemic equilibrium point cannot exactly classify the stability states of the COVID-19 dynamic system, it can be used to characterize how far the dynamic system is from the stable state. Table 2 listed the reproduction number 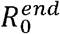 in all three steady periods across 50 states in the US, and showed that the number of states with 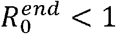 in the first, second and third steady period were 2, 4 and 19 states, respectively. Figure 3 showed Box plot for the reproduction number 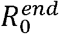 in three steady periods across 50 states in the US. We observed from Figure 3 that the median of 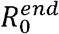 was down toward 1, which implied that the transmission dynamics of COVID-19 in the US were gradually toward stable states. A map of distributions of the reproduction number 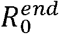 in three steady periods across 50 states in the US was plotted in Figure 4. As we can see from the third steady period in Figure 4, most states in the Middle East were expected to enter steady state in the recent COVID-19 wave in the US. Comparing Tables 1 and 2, we found that 16 states with 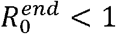 in the third steady period corresponded the states with decreasing order of the largest eigenvalues from the second steady period to the third steady period. This result demonstrated that both eigenvalues and the reproduction number in the steady periods characterized similar patterns of toward stability trend of COVID-19 across 50 states in the US.

**Table 2.** The reproduction number at the endemic equilibrium point in three steady periods across 50 states in the US.

| State | April - July, 2021 | March - May, 2022 | September - November, 2022 |
| --- | --- | --- | --- |
| AK | 1.4882 | 1.2717 | 1.6087 |
| AL | 1.6356 | 1.4859 | 1.4873 |
| AR | 1.3871 | 1.5872 | **0.7592** |
| AZ | 1.2515 | 1.4992 | **1.1714** |
| CA | 3.2000 | 1.4639 | **1.1679** |
| CO | 1.4619 | 0.9392 | 1.2262 |
| CT | 1.6514 | 1.1172 | 1.4204 |
| DE | 2.3482 | 1.4158 | **0.9796** |
| FL | 1.6243 | 1.3346 | 1.6129 |
| GA | 1.4301 | 1.3906 | 1.4375 |
| HI | 1.5330 | 1.2221 | 1.3806 |
| IA | 1.4979 | 2.3558 | 1.0906 |
| ID | 1.2902 | 1.5004 | **1.1492** |
| IL | 1.1010 | 1.2427 | **1.0303** |
| IN | 1.4808 | 2.0483 | **1.3011** |
| KS | 1.6181 | 1.5825 | **0.8463** |
| KY | 1.5401 | 1.1988 | **0.9389** |
| LA | 1.2269 | 1.5380 | 1.7963 |
| MA | 2.4008 | 1.0846 | **0.7365** |
| MD | 1.8112 | 1.4433 | **1.1016** |
| ME | 2.2400 | 1.4987 | **1.1074** |
| MI | 2.5159 | 1.2283 | **0.9385** |
| MN | 2.1400 | 1.6076 | **1.0471** |
| MO | 1.3783 | 1.3245 | **1.1722** |
| MS | 1.3353 | 1.7490 | 2.5765 |
| MT | 1.3814 | 1.9140 | 1.2367 |
| NC | 1.5884 | 0.8518 | 0.9769 |
| ND | 1.8152 | 1.6852 | **0.9715** |
| NE | 2.4089 | 2.8713 | 2.4377 |
| NH | 2.1694 | 1.1846 | **1.1882** |
| NJ | 1.8680 | **1.1596** | **1.0438** |
| NM | 1.2206 | **1.2357** | **1.0108** |
| NV | 0.9301 | 1.3862 | 1.5088 |
| NY | 1.8802 | 1.3264 | **0.9069** |
| OH | 1.6439 | 2.0173 | **0.8531** |
| OK | 1.6449 | 1.9003 | **1.0150** |
| OR | 1.4383 | **1.1184** | **1.0886** |
| PA | 2.4117 | 1.3496 | **0.9778** |
| PR | 4.1541 | 1.1200 | **0.9633** |
| RI | 1.2908 | 1.2471 | **0.8438** |
| SC | 1.5699 | 2.0826 | 1.0259 |
| SD | 1.7168 | 1.6564 | **0.9446** |
| TN | 1.6918 | 2.0071 | **0.9094** |
| TX | 1.2565 | 0.8002 | 1.5579 |
| UT | 1.2628 | 1.8566 | 1.4559 |
| VA | 2.3099 | 1.0337 | **0.8755** |
| VT | 1.7794 | 1.5289 | 1.1950 |
| WA | 1.4924 | 0.7499 | 0.9286 |
| WI | 1.5690 | 1.6867 | **0.9044** |
| WV | 1.8349 | 1.4470 | **0.8511** |
| WY | 0.7828 | 2.0850 | **0.9964** |

**Figure 3.**
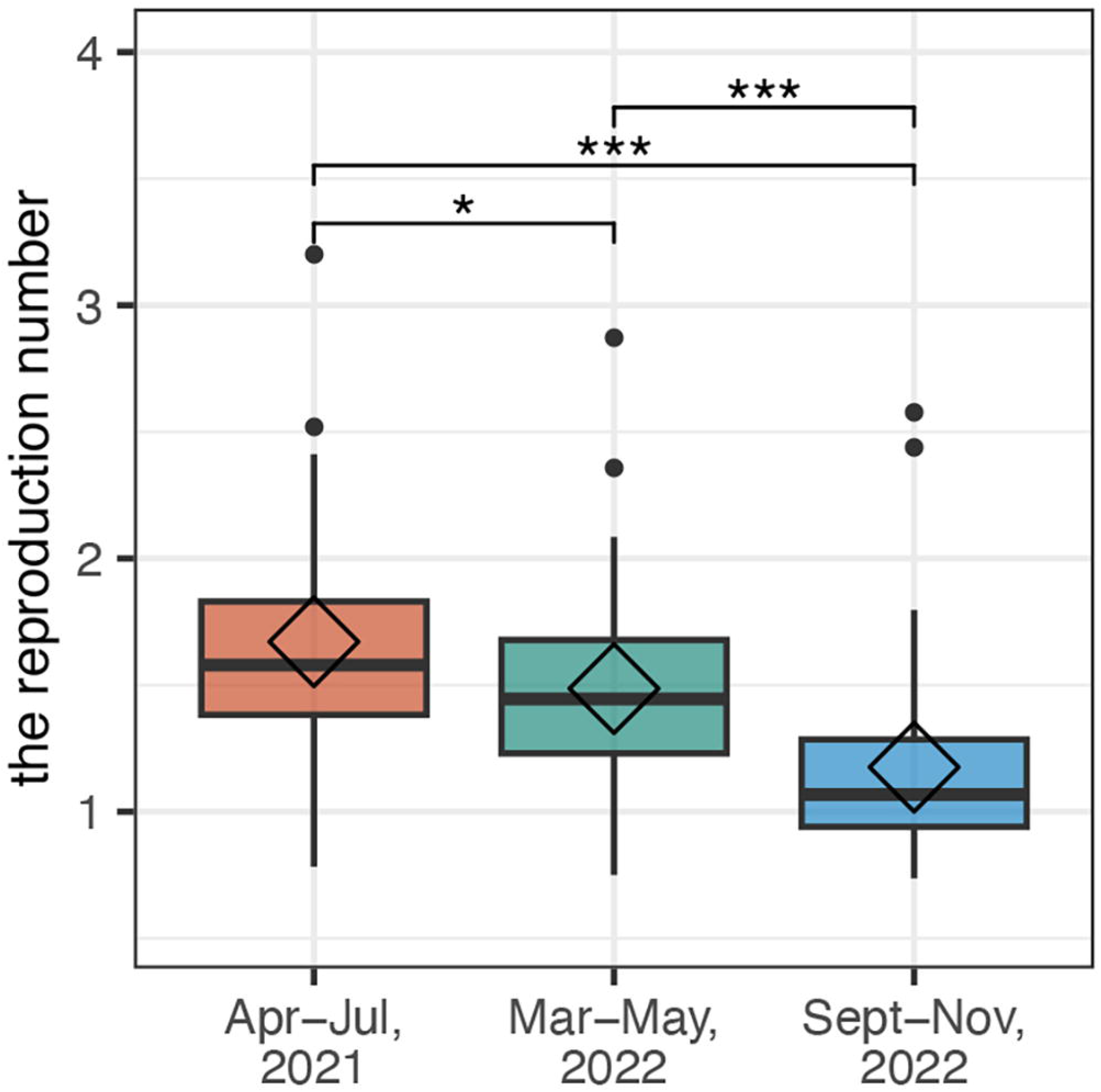
Box plot for the reproduction number 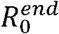 in three steady periods across 50 states in the US.

**Figure 4.**
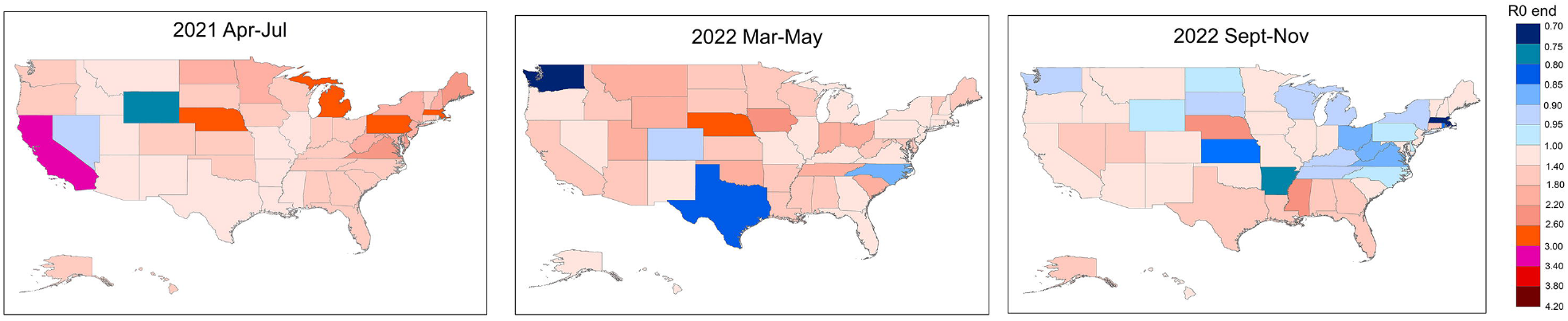
Map of distributions of the reproduction number 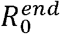 in three steady periods across 50 states in the US.

### 3.3. Impact of the parameters on the eigenvalues

To show the impact of the parameters *β, α, σ, γ*, and *δ* on the largest eigenvalue of the COVID-19 dynamics, we plotted Figure 5 to show the relationship between the parameters and the largest eigenvalue of the COVID-19 dynamic systems. We observed that the largest eigenvalue was positively correlated with the parameters *β*, or *σ*, or *γ*, or *δ* (during the second and third steady periods) and negatively correlated with the parameter *α*. This implied that to reduce the largest eigenvalue value, we needed to reduce the transmission-related parameters *β, γ* and *δ* or increase the vaccination rate *α* and the vaccine effeteness 1 − *σ*.

**Figure 5.**
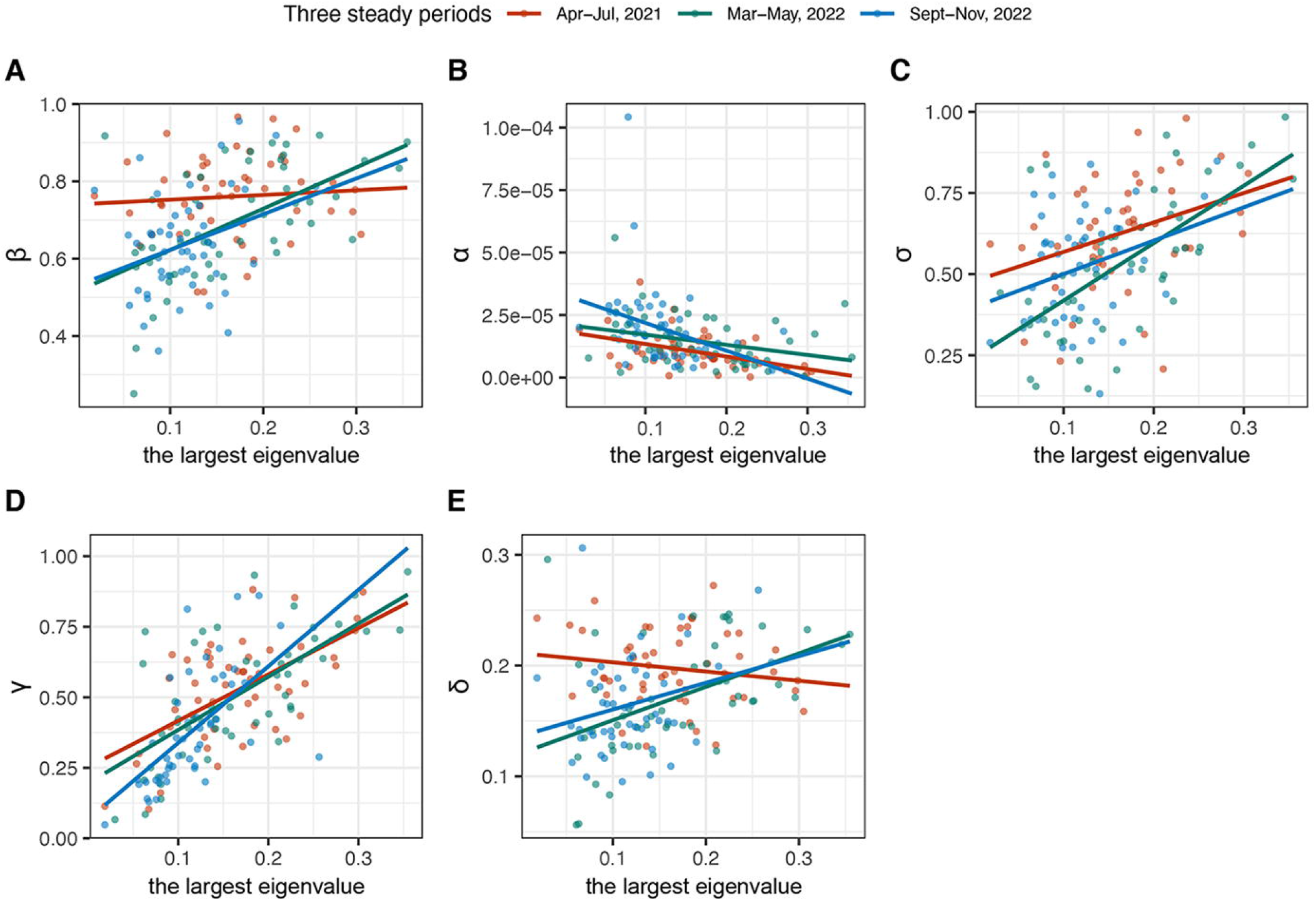
The relationship between the parameters and the largest eigenvalue of the COVID-19 dynamic systems. (A) The relationship between parameter β and the largest eigenvalue, (B) the relationship between parameter *α* and the largest eigenvalue, (C) the relationship between parameter *σ* and the largest eigenvalue, (D) the relationship between parameter *γ* and the largest eigenvalue, and (E) the relationship between parameter *δ* and the largest eigenvalue. The stability region was represented in blue color and instability region was represented in red color.

### 3.4. Approximate attractor analysis and impact of the parameters on stability

An attractor is defined as a set of states toward which a system attempts to return after perturbation.^17^ Instead of studying the attractor of the state variables *I, V, S, E, R* in the system, we studied the regions of the parameters that determine the stability of the COVID-19 dynamics and the impact of changing parameters on the stability. Our model included five estimated parameters, and it is difficult to present and visualize the stability region consisting of all the five parameters simultaneously.

Since the vaccination rate *α* and vaccine inefficiency rate *σ* jointly contribute to the transmission dynamics of COVID-19, we jointly investigated the impact of parameters *α* and *σ* together on the stability of COVID-19 dynamics. Figure S2 showed the impact of simultaneously changing parameters *α* and *σ* while keeping the current values of other parameters unchanged on the stability of COVID-19 dynamics in the steady periods (September – November 2022) across 50 states in the US. The stability region of the vaccination rate *α* and vaccine inefficiency rate *σ*is defined as the potential values of the parameters *α* and *σ* under which and current values of other parameters *β, γ*, and *δ* the COVID-19 dynamic system is stable. The other potential values of the parameters *α* and *σ* formed instability region under which and current values of other parameters *β, γ*, and *δ*, the COVID-19 dynamic system is unstable. The stability region was represented in blue color and instability region was represented in red color. Moving vertical axis up indicated an increase of the vaccination rate *α* and moving the horizontal axis left indicated a decrease of the inefficiency rate *σ* of vaccine or increase of the effect of vaccine. We observed from Figure S2 that among 50 states, Arkansas(AR), Kentucky (KY), Idaho (ID), Georgia (GA) had the biggest stability regions of *α* and *σ*, while Alaska (AK), Florida (FL), Louisiana (LA), Connecticut (CT) and Texas (TX) had the smallest stability regions of *α* and *σ*. Figure S2 provided valuable information that when the vaccine efficiency is low, we can increase the vaccination rate; or when the vaccination rate is low, we can increase the vaccine efficiency to ensure stability.

Since the transmission parameters *γ* and *δ* jointly contribute to the transmission dynamics of COVIFD-19, again, we investigated the impact of both parameters *γ* and *δ* on the stability of COVID-19 dynamics. Figure S3 showed the impact of simultaneously changing parameters *γ* and *δ* while keeping the current values of other parameters unchanged on the stability of COVID-19 dynamics in the steady periods (September – November 2022) across 50 states in the US. We observed that increasing the recovery rate *δ* (assuming no re-infection) and decreasing the incubation rate *γ* will move the dynamic system of COVID-19 toward the stability region. We observed from Figure S3 that among 50 states, Colorado (CO), Connecticut (CT), Montana (MT), New Jersey (NJ) and New York (NY) had the biggest stability regions of *γ*and *δ*, while Arkansas (AR), Iowa (IA), Nebraska (NE), Nevada (NV), Texas (TX) and Washington (WA) had the smallest stability regions of *γ* and *δ*.

Similarly, we plotted Figures S4 and S5 showing the impact of simultaneously changing parameters *γ* and *β*, and *δ* while keeping the current values of other parameters unchanged on the stability of COVID-19 dynamics in the steady periods (September – November 2022) across 50 states in the US, respectively. Figure S4 showed that simultaneously increasing parameter *β* and decreasing parameter *γ* will drive the dynamic system of COVID-19 toward stability region. Figure S4 also showed that the parameter *γ* is more important to push the dynamics of COVID-19 toward stability states than the parameter *β* and the stability region of the parameters *γ* and *β* is smaller than that of parameters *γ* and *δ*. Figure S5 showed that simultaneously decreasing the transmission parameter *β* and increasing the recovery rate *δ* while keeping the current values of other parameters unchanged will move the dynamic system of COVID-19 toward stability region. In Figure 5, we observed that the transmission parameter *β* is more important in driving the dynamics of COVID-19 toward stability states than the recovery rate *δ*.

## 4. Conclusions

Currently, the most popular and urgent question is when will the COVID-19 pandemic end.^18^ Some scientists optimistically predict that “the COVID19 pandemic could terminate in 2022”.^19^ Other scientists warn that COVID-19 pandemic is far from over.^20-22^ In this paper, we develop stability of transmission dynamics of COVID-19 as a measure to assess whether the COVID-19 pandemic is evolved to the endemic.

There is no consensus about definition of pandemic.^23^ Pandemic and endemic are concepts of systems dynamics. We interpretate the endemic as “stable states” of the COVID-19 dynamic system. Protecting people from infection of COVID-19, in general, depends on vaccination and natural immunity. Due to lack of natural immunity data, we develop SEIR mathematical model with vaccination only included to study the stability of the COVID-19 dynamics. We formulate the objective function for ensuring the steady states of dynamics which is then used to estimate the parameters underlying the current transmission dynamics of COVID-19. We derived eigenequations and found eigenvalues as well as reproduction number *R*_0_ underlying disease-free equilibrium point and 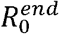 underlying the endemic equilibrium point for assessing the stability of the COVID-19 dynamics under these two types of equilibrium points. We obtained two remarkable conclusions. First conclusion is that all 50 states in the US in all three steady periods were in unstable states. Second conclusion is, however, COVID-19 dynamics of all 50 states in the US were toward stable states. Real data analysis showed that three transition related parameters *β, γ* and *δ* decreased while the vaccination rate *α* and vaccine efficiency 1 − *σ* increased when we went from the first steady period through the second steady period to the third steady period. Again, the results also showed that the largest eigenvalue of the stability analysis matrix and the reproduction number 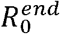 in three steady periods decreased when we went from the first steady period through the second steady period to the third steady period.

Both vaccination and non-pharmaceutical interventions (NPIs) for COVID-19 are important tools for controlling the spread of COVID-19. In theory, we can search the space of vaccination related parameters *α, σ* and NPI related parameters *β, γ, δ*, to form the stability region of the parameters in the steady state period. In this report, we calculated the stability regions of *α, σ*, and *β, γ, δ*, which provided information on how to changing COVID-19 from pandemic to endemic. Improving vaccine efficiency (for example, using mucosal vaccines) and increasing vaccination rate with appropriate NPIs will change COVID-19 from pandemic to endemic.

Virus evolution is uncertain. Transition dynamics of COVID-19 governed by virus variant evolution, emergence of new variants, vaccination and unknown natural immunity in the host is complex and uncertain. There is not a simple and direct path from pandemic to endemic. As editorial of “nature” pointed out “there won’t be a single ‘exit’ wave to mark the lifting of pandemic restrictions. Further waves of infection and death are likely to follow, either from new variants that arise in the population, or from variants imported as the country opens its borders to visitors”,^24^ COVID-19 in the US has experienced five major waves and may have more waves. Good news is that COVID-19 in the US is getting closer and closer to endemic.

## Data Availability

All data produced are available online at https://github.com/nytimes/covid-19-data
https://ourworldindata.org/us-states-vaccinations.

https://ourworldindata.org/us-states-vaccinations.

## Acknowledgments

Zixin Hu in this study was partially supported by funding from the National Natural Science Foundation of China (3210040426 to Z.H.), the Shanghai Rising-Star Program (21QB1400900 to Z.H.), and was also partly supported by a grant from the major project of Study on Pathogenesis and Epidemic Prevention Technology System (2021YFC2302500) by the Ministry of Science and Technology of China.

## Author Contribution Statement

Perform data analysis and write paper: Zixin Hu, Xiaoxi Hu, Kai Zhang

Provide Data and Result Interpretation: Tao Xu, Jinying Zhao

Result Intepretation: Henry H Lu

Design project: Li Jin, Eric Boerwinkle

Design project and write paper: Momiao Xiong

## Figure S Legend

**Figure S1**. Three steady state periods (April – July, 2021; March – May, 2022; and September – November, 2022) are selected from January 12, 2021 to December 12, 2022. Three periods are represented in red, green and blue colors.

**Figure S2**. Impact of simultaneously changing parameters *α* and *σ* while keeping the current values of other parameters unchanged on the stability of COVID-19 dynamics in the steady periods (September – November 2022) across 50 states in the US. Vertical axis represented *α* and horizontal axis *σ*.

**Figure S3**. Impact of simultaneously changing parameters *γ* and *δ* while keeping the current values of other parameters unchanged on the stability of COVID-19 dynamics in the steady periods (September – November 2022) across 50 states in the US. Vertical axis represented *γ* and horizontal axis *δ*.

**Figure S4**. Impact of simultaneously changing parameters *γ* and *β* while keeping the current values of other parameters unchanged on the stability of COVID-19 dynamics in the steady periods (September – November 2022) across 50 states in the US. Vertical axis represented *γ* and horizontal axis *β*.

**Figure S5**. Impact of simultaneously changing parameters *δ* and *β* while keeping the current values of other parameters unchanged on the stability of COVID-19 dynamics in the steady periods (September – November 2022) across 50 states in the US. Vertical axis represented *β* and horizontal axis *δ*.

**Figure S6**. Impact of simultaneously changing parameters *β, γ* and *δ* while keeping the current values of other parameters unchanged on the stability of COVID-19 dynamics in the steady periods (September – November 2022). Axis *x* represented parameter *β*, axis *y* represented parameter *δ* and axis *z* represented parameter *γ*. (A) NY, the state with the largest stability region, (B) CT, the state with the second largest stability region, (C) TX, the state with the smallest stability region, and (D) NE, the state with the second smallest stability region.

## REFERENCES

1. COVID Data Tracker Weekly Review – CDC. https://www.cdc.gov/coronavirus/2019ncov/covid-data/covidview/index.html.

2. del Rio C, Malan PN. COVID-19 in 2022—The beginning of the end or the end of the beginning? JAMA. 2022;327(24):2389–2390.

3. Wang et al., 2023, Alarming antibody evasion properties of rising SARSCoV-2 BQ and XBB subvariants. Cell 186, 1–8.

4. WHO Coronavirus (COVID-19) Dashboard. https://covid19.who.int/.

5. Hsiang S, Allen D,Annan-Phan S, et al. The effect of large-scale anti-contagion policies on the COVID-19 pandemic. medRxiv 2020.03.22.20040642. https://www.medrxiv.org/content/10.1101/2020.03.22.20040642v4.full.pdf.

6. Li Q, Guan X, Wu P et al. Early transmission dynamics in Wuhan, China, of novel Coronavirus-infected pneumonia. N Engl J Med. 2020; 38:1199–1207.

7. Wu JT, Leung K, Leung GM. Nowcasting and forecasting the potential domestic and international spread of the 2019-nCoV outbreak originating in Wuhan, China: a modelling study. Lancet. 2020; 395:689–697.

8. Zhao S. Musa SS, Lin Q, et al. Estimating the unreported number of novel Coronavirus (2019-nCoV) cases in China in the first half of January 2020: A data-driven modelling analysis of the early outbreak. J Clin Med. 2020; 9: pii: E388. doi: 10.3390/jcm9020388.

9. Kucharski A, Russell T, Diamond C, Liu Y. CMMID nCoV working group, Edmunds J, Funk S, Eggo R. Analysis and projections of transmission dynamics of nCoV in Wuhan. (2020) https://cmmid.github.io/ncov/wuhan_early_dynamics/index.html.

10. Tuite AR, Fisman DN. Reporting, epidemic growth, and reproduction numbers for the 2019 novel coronavirus (2019-nCoV) epidemic. Ann Intern Med. 2020; 172:567–568.

11. Hellewell J, Abbott S, Gimma A, et al. Centre for the Mathematical Modelling of Infectious Diseases COVID-19 Working Group, Funk S1, Eggo RM2. Feasibility of controlling COVID-19 outbreaks by isolation of cases and contacts. Lancet Glob Health 2020; 8:e488–e496.

12. Ghostine R, Gharamti M, Hassrouny S, Hoteit I. An Extended SEIR Model with Vaccination for Forecasting the COVID-19 Pandemic in Saudi Arabia Using an Ensemble Kalman Filter. Mathematics 2021; 9: 636.

13. Viana J, van Dorp CH, Nunes A, et al. Controlling the pandemic during the SARS-CoV-2 vaccination rollout. Nat Commun. 2021;12:3674.

14. Brand SPC, Ojal J, Rabia Aziza R, et al. COVID-19 transmission dynamics underlying epidemic waves in Kenya. Science. 2021;eabk0414.

15. Sun YJ, Wu YB, Wang CC. Existence and uniqueness of the exponentially stable limit cycle for a class of nonlinear systems via time-domain approach with differential inequality. Journal of Applied Mathematics. 2013; Article ID 712932.

16. van den Driessche, P. Reproduction numbers of infectious disease models. Infectious Disease Modelling. 2017; 2(3): 288–303.

17. Attractor. https://en.wikipedia.org/wiki/Attractor.

18. Charumilind S, et al. When will the COVID-19 pandemic end? https://www.mckinsey.com/industries/healthcare-systems-and-services/our-insights/when-will-the-covid-19-pandemic-end.

19. Chen JM. Novel statistics predict the COVID-19 pandemic could terminate in 2022. J Med Virol. 2022; 94(6):2845–2848.

20. Editorial. There’s no room for COVID complacency in 2023. Nature 2023; 613: 7.

21. Editorial. The COVID-19 pandemic in 2023: far from over. The Lancet 2023; 401.

22. Robertson D. COVID in 2023 and beyond – why virus trends are more difficult to predict three years on. https://theconversation.com/covid-in-2023-and-beyond-why-virus-trends-are-more-difficult-to-predict-three-years-on-196170.

23. Shmerling RH. Is the COVID-19 pandemic over, or not? Harvard Health Publishing. October 26, 2022.

24. Moore S, Hill EM, Tildesley MJ, Dyson L, Keeling MJ. Vaccination and non-pharmaceutical interventions for COVID-19: a mathematical modelling study. The LANCET. 21 (6): 793–802.

