## Supplemental for "The Equilibrium and Pandemic Waves of COVID-19 in the US"

**Supplementary Material**

**Supplementary Note**

**Supplementary Figures: 6**

### Supplementary Note

### 1. SEIRV compartment model for COVID-19 transmission dynamics

We constructed a compartmental model based on a deterministic system of nonlinear differential equations that considers vaccination. The architecture of the SEIRV is shown in Figure 1.

Let $S(t), E(t), I(t), R(t)$ and $V(t)$ be the number of susceptible, exposed, infectious, recovered and vaccinated individuals at time $t$.[9] Define the parameters as follows.

$\Lambda$: the new birth and new residents per unit of time,

$\beta$: transmission rate divided by the total population $N$,

$\alpha$: vaccination rate,

$\mu$: natural death rate,

$\gamma$: incubation rate by which the exposed individual develops symptoms,

$\delta$: the probability of the recovery or death,

$\sigma$: vaccine inefficiency.

Now we define a set of nonlinear ordinary differential equations as a mathematic model for the transmission dynamics of COVID-19:

$$\frac{dS(t)}{dt} = \Lambda - \beta S(t)I(t) - \alpha S(t) - \mu S(t)\ , \quad \text{(S1)}$$

$$\frac{dE(t)}{dt} = \beta S(t)I(t) - \gamma E(t) + \sigma\beta I(t)V(t) - \mu E(t), \quad \text{(S2)}$$

$$\frac{dI(t)}{dt} = \gamma E(t) - \delta I(t) - \mu I(t), \quad \text{(S3)}$$

$$\frac{dR(t)}{dt} = \delta I(t) - \mu R(t), \quad \text{(S4)}$$

$$\frac{dV(t)}{dt} = \alpha S(t) - \sigma\beta I(t)V(t) - \mu V(t). \quad \text{(S5)}$$

Let $N(t) = S(t) + E(t) + I(t) + R(t) + V(t)$ be the total population size.

**2. Non-negativity and boundedness of the solutions**

In this section we show the non-negativity and boundedness of the solutions of the nonlinear differential equations (S1-S5). Assume that all initial values are non-negative. Since $\Lambda \geq 0$, it follows from equation S(1) that

$$\frac{dS}{dt} \geq -(\beta I(t) + \alpha + \mu)S(t), \quad \text{(S6)}$$

which implies that

$$S(t) \geq S(0)e^{-\int_0^t (\beta I(\tau)+\alpha+\mu)d\tau} \geq 0 \, . \quad \text{(S7)}$$

By the similar arguments, we can show that all $E(t), I(t), R(t)$ and $V(t)$ are non-negative. For example, using equation (S5) and (S7), we obtain

$$\frac{dV}{dt} \geq -(\sigma\beta I(t) + \mu)V(t)$$

$$\geq V(0)e^{-\int_0^t (\sigma\beta I(\tau)+\mu)d\tau} \geq 0 \, .$$

Now we show that all solutions are bounded by $\frac{\Lambda}{\mu}$ if we assume that the initial value $N(0)$ is also bounded by $\frac{\Lambda}{\mu}$, i.e., $N(0) \leq \frac{\Lambda}{\mu}$. Summarizing equations (S1)-(S5), we obtain

$$\frac{dN}{dt} = \Lambda - \mu N(t) \, . \quad \text{(S8)}$$

Solving equation (S30) for $N(t)$, we obtain

$$N(t) = \frac{\Lambda}{\mu} + \left(N(0) - \frac{\Lambda}{\mu}\right) e^{-\mu t} \, . \tag{S9}$$

If $N(0) \leq \frac{\Lambda}{\mu}$ then $\left(N(0) - \frac{\Lambda}{\mu}\right) e^{-\mu t} \leq 0$, which implies

$$N(t) \leq \frac{\Lambda}{\mu} \, . \tag{S10}$$

Equation (S10) indicates that all variables $S(t), E(t), I(t), R(t), V(t)$ are in the region $\Omega$:

$$\Omega = \{\mathrm{N(t) = S(t) + E(t) + I(t) + R(t) + V(t)} | 0 \leq N(t) \leq \frac{\Lambda}{\mu}\} \, . \tag{S11}$$

**3. Steady state analysis of COVID-19 transmission dynamical systems**

Steady state of a dynamic system is the state that will not change without external excitation. Steady state analysis includes the basic reproduction number, critical epidemic equilibrium points, and endemic equilibrium points of the COVID-19 model.

**3.1 Reproduction number**

Assume that the compartments are divided into two sets. The first set contains infected individuals $(E, I)$. The second set consists of remaining individuals $(S, R, V)$. Let

$$X(t) = \begin{bmatrix} x_1(t) \\ x_2(t) \end{bmatrix} = \begin{bmatrix} E(t) \\ I(t) \end{bmatrix} \, .$$

Define

$$\frac{dx_i(t)}{dt} = \mathcal{F}_i(X) - \mathcal{V}_i(x), i = 1, 2, \tag{S12}$$

where $\mathcal{F}_i$is the rate of appearance of new infections in compartment $i$ and $\mathcal{V}_i(x)$ includes the rate of transitions between compartment $i$ and other infected compartments.

Recall that

$$\frac{dx}{dt} = \begin{bmatrix} \frac{dE}{dt} \\ \frac{dI}{dt} \end{bmatrix} = \begin{bmatrix} -(\gamma+\mu)E(t) + \beta\big(S(t)+\sigma V(t)\big)I(t) \\ \gamma E(t) - (\delta+\mu)I(t) \end{bmatrix}. \qquad \text{(S13)}$$

The rate of appearance of new infections in compartment $E$ is $\beta\big(S(t)+\sigma V(t)\big)I(t)$ and the rate of appearance of new infections in compartment $I(t)$ is zero. The rate of transitions between compartment $E(t)$ and other infected compartments is $-(\gamma+\mu)E(t)$ and the rate of transitions between compartment $I(t)$ and other infected compartments includes $\gamma E(t) - (\delta+\mu)I(t)$. Thus, equation (S13) can be written as

$$\frac{dX}{dt} = \begin{bmatrix} \beta\big(S(t)+\sigma V(t)\big)I(t) \\ 0 \end{bmatrix} - \begin{bmatrix} (\gamma+\mu)E(t) \\ -\gamma E(t) + (\delta+\mu)I(t) \end{bmatrix}$$

$$= \mathcal{F}(X) - \mathcal{V}(x)\,. \qquad \text{(S14)}$$

Let

$F(X) = \left[\frac{\partial\mathcal{F}(X)}{\partial X^T}\right]$ and $V(X) = \left[\frac{\partial\mathcal{V}(x)}{\partial X^T}\right]$.

Then, using equation (S14), we obtain

$$F(X) = \begin{bmatrix} 0 & \beta\big(S(t)+\sigma V(t)\big) \\ 0 & 0 \end{bmatrix}, \qquad \text{(S15)}$$

$$V(X) = \begin{bmatrix} (\gamma+\mu) & 0 \\ -\gamma & (\delta+\mu) \end{bmatrix}, V^{-1}(X) = \frac{1}{(\gamma+\mu)(\delta+\mu)} \begin{bmatrix} \delta+\mu & 0 \\ \gamma & \gamma+\mu \end{bmatrix}. \qquad \text{(S16)}$$

Using next generation matrix method (van den Driessche 2017) to calculate the basic reproduction number $R_0$, we first calculate

$$F(X)V^{-1}(X) = \frac{1}{(\gamma+\mu)(\delta+\mu)} \begin{bmatrix} \gamma\beta\big(S(t)+\sigma V(t)\big) & (\gamma+\mu)\beta\big(S(t)+\sigma V(t)\big) \\ 0 & 0 \end{bmatrix}. \qquad \text{(S17)}$$

It is easy to see that the eigenvalues of the matrix $F(X)V^{-1}(X)$ are given by

$$\lambda_1 = \frac{\gamma\beta\left(S(t)+\sigma V(t)\right)}{(\gamma+\mu)(\delta+\mu)}\ , \lambda_2 = 0\ .$$

The basic reproduction number $R_0$ is defined as the  the spectral radius of the next-generation matrix $F(X)V^{-1}(X)$:

$$R_0 = \frac{\gamma\beta\left(S(t)+\sigma V(t)\right)}{(\gamma+\mu)(\delta+\mu)}\ . \qquad \text{(S18)}$$

**3. 2. Critical Point and equilibrium of COVID-19 dynamic systems**

Critical transition of COVID-19 dynamic systems occurs when restrictions on travel, social gathering and meeting, open of school, mandatory wearing of masks are lifted or

poorly adhered to, new variants with strong transmission rates, successfully invade. COVID-19 dynamic system has multiple stable equilibrium points. There  must also be unstable equilibrium points between the stable points in the COVID-19 transmission dynamics.  Multistability determines the trend of COVID-19 transmission dynamics and provide information on designing public health intervention measures to eradicate CIVID-19 outbreaks. In this section we apply the Jacobian matrix-based linear stability analysis  to continuous-time nonlinear dynamical systems. For convenience of discussion, nonlinear differential equations (S1-S5)  is rewritten as:

$$\frac{dx}{dt} = f(x)\ . \qquad \text{(S19)}$$

A critical point of the system is a point $(x_*)$ such that:

$$f(x_*) = 0\ . \qquad \text{(S20)}$$

In other words,  a constant solution $x_*$ at a critical point satisfies both $\frac{dx_*}{dt} = 0$ and $f(x_*) = 0$ and hence

$$\frac{dx_*}{dt} = f(x_*)$$

The solution that begins with critical point $x_*$ and then just stays at the critical point $x_*$. Therefore, the critical point $x_*$ is also called an equilibrium solution. The critical point (equilibrium) can be found by setting the left sides of differential equations (S1-S5) to be zero. Therefore, the critical points of COVID-19 dynamic system are defined by the following five algebraic equations.

$$\Lambda - \beta S_* I_* - (\alpha + \mu) S_* = 0, \quad \text{(S21)}$$

$$\beta S_* I_* + \sigma\beta I_* V_* - (\gamma + \mu) E_* = 0\ , \quad \text{(S22)}$$

$$\gamma E_* - (\delta + \mu) I_* = 0\ , \quad \text{(S23)}$$

$$\delta I_* - \mu R_* = 0\ , \quad \text{(S24)}$$

$$\alpha S_* - \sigma\beta I_* V_* - \mu V_* = 0\ . \quad \text{(S25)}$$

Now we find critical points by solving equations (S21-S25).

Solving equation (S23) yields

$$E_* = \frac{\delta+\mu}{\gamma} I_*. \quad \text{(S26)}$$

Substituting equation (S26) into equation (S22), we obtain

$\beta S_* I_* + \sigma\beta I_* V_* - (\gamma + \mu)\frac{\delta+\mu}{\gamma} I_* = 0$ , which is reduced to

$$I_* \left(\beta S_* + \sigma\beta V_* - \frac{(\gamma+\mu)(\delta+\mu)}{\gamma}\right) = 0. \quad \text{(S27)}$$

Solving equation (S27), we obtain two solutions:

(1) $I_* = 0$ , (S28)

(2) $I_* \neq 0, \beta S_* + \sigma\beta V_* - \frac{(\gamma+\mu)(\delta+\mu)}{\gamma} = 0.$ (S29)

Since the number of new cases $I_*$ determines the disease status, $I_* = 0$ indicates the disease-free, and hence critical point $I_* = 0$ is called *disease-free* critical point or *disease free* equilibrium. The second critical point defined in equation (S29) is called the *endemic* critical point or equilibrium.

In scenario (1), we obtain

$E_* = 0$ (equation S26), $R_* = 0$ (equation S24), $S_* = \frac{\Lambda}{\alpha+\mu}$ (equation S21), and $V_* = \frac{\alpha\Lambda}{\mu(\alpha+\mu)}$ (equations S21 and S25). In summary, *the disease-free critical point* is

$$I_* = 0, E_* = 0, R_* = 0, S_* = \frac{\Lambda}{\alpha+\mu}, V_* = \frac{\alpha\Lambda}{\mu(\alpha+\mu)}, R_0 = \frac{\gamma\beta\Lambda(\mu+\alpha\sigma)}{\mu(\gamma+\mu)(\delta+\mu)(\alpha+\mu)}. \quad \text{(S30)}$$

Now we discuss scenario (2) $I_* \neq 0$.

Let

$$a = \frac{\sigma(\gamma+\mu)(\delta+\mu)\beta^2}{\mu}$$

$$b = \frac{(\gamma+\mu)(\delta+\mu)\beta}{\gamma\mu}[\mu + (\alpha+\mu)\sigma] - \frac{\Lambda\sigma\beta^2}{\mu}$$

$$c = \frac{(\gamma+\mu)(\delta+\mu)(\alpha+\mu)}{\gamma} + \beta[\Lambda\sigma - \Lambda - \frac{\alpha+\mu}{\mu}\Lambda]$$

$$d = (\Lambda - \mu)(\alpha+\mu) ,$$

$$F(\Lambda, \mu, \delta, \gamma, \beta, \alpha, \sigma) = aI^3 + bI^2 + cI + d .$$

The parameters are estimated by minimizing

$$\min_{\Lambda,\mu,\delta,\gamma,\beta,\alpha,\sigma} F(\Lambda,\mu,\delta,\gamma,\beta,\alpha,\sigma)^2.$$

Solving equation (S21), we obtain

$$S_* = \frac{\Lambda}{\alpha+\mu+\beta I_*} . \quad \text{(S31)}$$

Substituting equation (S31) into equation (S25) yields

$$V_* = \frac{\alpha}{\mu+\sigma\beta I_*} S_* = \frac{\alpha\Lambda}{(\mu+\sigma\beta I_*)(\alpha+\mu+\beta I_*)} . \quad \text{(S32)}$$

Solving equation (23) yields

$$E_* = \frac{\delta+\mu}{\gamma} I_* . \quad \text{(S33)}$$

Solving equation (S24) yields

$$R_* = \frac{\delta}{\mu} I_* . \quad \text{(S34)}$$

Substituting equations (S32-S34) into equation (S22) yields

$$aI_*^2 + bI_* + c = 0 , \quad \text{(S35)}$$

where

$$a = \frac{(\gamma+\mu)(\delta+\mu)\sigma\beta^2}{\gamma} , \quad \text{(S36)}$$

$$b = \frac{(\gamma+\mu)(\delta+\mu)[\mu\beta+(\alpha+\mu)\sigma\beta]}{\gamma} - \sigma\beta^2\Lambda , \quad \text{(S37)}$$

$$c = \frac{\mu(\gamma+\mu)(\delta+\mu)(\alpha+\mu)}{\gamma} - \beta\Lambda(\mu+\alpha\sigma) . \quad \text{(S38)}$$

Solving quadratic equation (S35) for $I_*$, we obtain

$$I_* = \frac{-b \pm \sqrt{b^2 - 4ac}}{2a} \, . \qquad \text{(S39)}$$

Since $I_* \geq 0$, then we obtain the equilibrium point:

$$I_* = \frac{-b + \sqrt{b^2 - 4ac}}{2a} \, , \qquad \text{(S40)}$$

where

$c < 0.$

If we assume that natural death rate $\mu$ is zero, then we obtain

$$a = \delta\sigma\beta^2, b = \sigma\beta(\alpha\delta - \beta\Lambda) \, , c = -\alpha\beta\sigma\Lambda. \qquad \text{(S41)}$$

Substituting equation (S41) into equation (S40), we obtain

$$I_* = \frac{\Lambda}{\delta} \, . \qquad \text{(S42)}$$

After $I_*$ is found, we can obtain

$S_* = \frac{\Lambda}{\alpha + \mu + \beta I_*}$ (equation S21),

$V_* = \frac{\alpha\Lambda}{(\mu + \sigma\beta I_*)(\alpha + \mu + \beta I_*)}$ (equation S32),

$E_* = \frac{\delta + \mu}{\gamma} I_*$ (equation S23)

$R_* = \frac{\delta}{\mu} I_*$ (equation S24) (S43)

In summary, we obtain two equilibrium points:

(1) Disease free critical (equilibrium) point:

$$I_* = 0, E_* = 0, R_* = 0, S_* = \frac{\Lambda}{\alpha+\mu}, V_* = \frac{\alpha\Lambda}{\mu(\alpha+\mu)}, R_0^{free} = \frac{\gamma\beta\Lambda(\mu+\alpha\sigma)}{\mu(\gamma+\mu)(\delta+\mu)(\alpha+\mu)}. \quad \text{(S44)}$$

(2) Endemic critical (equilibrium) point:

$$I_* = \frac{-b+\sqrt{b^2-4ac}}{2a},$$

$$a = \frac{(\gamma+\mu)(\delta+\mu)\sigma\beta^2}{\gamma}, b = \frac{(\gamma+\mu)(\delta+\mu)[\mu\beta+(\alpha+\mu)\sigma\beta]}{\gamma} - \sigma\beta^2\Lambda,$$

$$c = \frac{\mu(\gamma+\mu)(\delta+\mu)(\alpha+\mu)}{\gamma} - \beta\Lambda(\mu+\alpha\sigma),$$

$$S_* = \frac{\Lambda}{\alpha+\mu+\beta I_*}, \; V_* = \frac{\alpha\Lambda}{(\mu+\sigma\beta I_*)(\alpha+\mu+\beta I_*)}, E_* = \frac{\delta+\mu}{\gamma} I_*, \; R_* = \frac{\delta}{\mu} I_* .$$

$$R_0^{end} = \frac{\gamma\beta\Lambda}{(\gamma+\mu)(\delta+\mu)(\alpha+\mu+\beta I_*)}\left[1 + \frac{\alpha\sigma}{(\mu+\sigma\beta I_*)}\right] \quad \text{(S45)}$$

#### 3.3. Classification of Critical Points

Stability analysis for the general nonlinear dynamic systems is complicated. In this paper, we will focus on isolated critical point and almost linear systems. If there is only critical point in its neighborhood, then this critical point is called an isolated critical point. A system is called almost linear at a critical point if the Jacobian matrix of linearized system at an isolated critical point is invertible. Let $x_*$ be an isolated critical point. Assume that the Jacobian matrix of the nonlinear dynamic system (S19 ) is invertible. Consider almost linear system of the nonlinear system at the isolated critical point:

$$\frac{dx}{dt} \approx \frac{\partial f}{\partial x^T}\Big|_{x=x_*} (x - x_*) . \quad \text{(S46)}$$

Denote the Jacobian matrix at the critical point $x_*$ as $J = \frac{\partial f}{\partial x^T}\Big|_{x=x_*}$. Using equations (S1-S5), we obtain

$$J = \begin{bmatrix} -\varepsilon_1 & 0 & -\beta S_* & 0 & 0 \\ \beta I_* & -\varepsilon_2 & \beta(S_* + \sigma V_*) & 0 & \sigma\beta I_* \\ 0 & \gamma & -\varepsilon_3 & 0 & 0 \\ 0 & 0 & \delta & -\mu & 0 \\ \alpha & 0 & -\sigma\beta V_* & 0 & -\varepsilon_4 \end{bmatrix}, \quad \text{(S47)}$$

where

$$\varepsilon_1 = \alpha + \mu + \beta I_*, \varepsilon_2 = \gamma + \mu, \varepsilon_3 = \delta + \mu \text{ and } \varepsilon_4 = \mu + \sigma\beta I_*. \quad \text{(S48)}$$

Once the Jacobian matrix is calculated, we then calculate its eigenvalues and classify the critical points. We first consider disease-free critical point.

#### 3.3.1 Disease-free critical point

Substituting equation (S44) into equation (S48), we obtain

$$\varepsilon_1^0 = \alpha + \mu, \varepsilon_2 = \gamma + \mu, \varepsilon_3 = \delta + \mu \text{ and } \varepsilon_4^0 = \mu. \quad \text{(S49)}$$

Again, substituting equation (S49) into equation (S47) yields the Jacobian matrix at the disease-fee critical point:

$$J^0 = \begin{bmatrix} -\varepsilon_1^0 & 0 & -\beta S_* & 0 & 0 \\ 0 & -\varepsilon_2 & \beta(S_* + \sigma V_*) & 0 & 0 \\ 0 & \gamma & -\varepsilon_3 & 0 & 0 \\ 0 & 0 & \delta & -\mu & 0 \\ \alpha & 0 & -\sigma\beta V_* & 0 & -\varepsilon_4^0 \end{bmatrix}. \quad \text{(S50)}$$

Its characteristic polynomial is given by

$$|\lambda I - J^0| = (\lambda + \varepsilon_1^0)(\lambda + \mu)(\lambda + \varepsilon_4^0)[(\lambda + \varepsilon_2)(\lambda + \varepsilon_3) - \gamma\beta(S_* + \sigma V_*)] . \quad \text{(S51)}$$

Using equation (S18), we obtain

$$\gamma\beta(S_* + \sigma V_*) = \varepsilon_2\varepsilon_3 R_0 , \quad \text{(S52)}$$

where $R_0$ is the basic reproduction number.

Substituting equation (S52) into equation (S51) yields

$$|\lambda I - J^0| = (\lambda + \varepsilon_1^0)(\lambda + \mu)(\lambda + \varepsilon_4^0)[\lambda^2 + (\varepsilon_2 + \varepsilon_3)\lambda + (1 - R_0)\varepsilon_2\varepsilon_3] \, . \tag{S53}$$

The solutions to characteristic equation (S53) are

$$\lambda_1 = -\varepsilon_1^0 = -(\alpha + \mu), \tag{S54}$$

$$\lambda_2 = \frac{-(\varepsilon_2+\varepsilon_3)+\sqrt{(\varepsilon_2-\varepsilon_3)^2+4\varepsilon_2\varepsilon_0 R_0}}{2} = \frac{-(\gamma+\delta+2\mu)+\sqrt{(\gamma-\delta)^2+4(\gamma+\mu)(\delta+\mu)R_0}}{2}, \tag{S55}$$

$$\lambda_3 = \frac{-(\varepsilon_2+\varepsilon_3)-\sqrt{(\varepsilon_2-\varepsilon_3)^2+4\varepsilon_2\varepsilon_0 R_0}}{2} = \frac{-(\gamma+\delta+2\mu)-\sqrt{(\gamma-\delta)^2+4(\gamma+\mu)(\delta+\mu)R_0}}{2}, \tag{S56}$$

$$\lambda_4 = \lambda_5 = -\varepsilon_4^0 = -\mu \, . \tag{S57}$$

Eigenvalues $\lambda_1$, $\lambda_3$, $\lambda_4$ and $\lambda_5$ are negative. Now we investigate $\lambda_2$. When $R_0 < 1$, then

$$(\gamma - \delta)^2 + 4(\gamma + \mu)(\delta + \mu)R_0 < (\gamma - \delta)^2 + 4(\gamma + \mu)(\delta + \mu)$$

$$= (\gamma + \delta + 2\mu)^2 \, . \tag{S58}$$

Using equation (S58), we obtain

$\sqrt{(\gamma - \delta)^2 + 4(\gamma + \mu)(\delta + \mu)R_0} < \gamma + \delta + 2\mu$, which implies that

$$-(\gamma + \delta + 2\mu) + \sqrt{(\gamma - \delta)^2 + 4(\gamma + \mu)(\delta + \mu)R_0} < 0 \, .$$

Therefore, $\lambda_2$ is negative. In other words, When $R_0 < 1$, then all solutions are negative, the system is locally asymptotically stable. By the similar arguments, when $R_0 = 1$, then $\lambda_2 = 0$ the system is unstable. When $R_0 > 1$, then $\lambda_2 > 0$. The system is unstable.

Therefore, **the disease-free critical point can be classified as three cases:**

(1) when $R_0 < 1$, the disease-free critical point is classified as a asymptotically stable node;

(2) when $R_0 = 1$, the disease-free critical point is classified as an unstable node; and

(3) when $R_0 > 1$, the disease-free critical point is classified as an unstable saddle point.

**3.3.2 Endemic equilibrium point**

Recall from the Jacobian matrix in equation (S47) that under the endemic equilibrium point condition , we have $I_* \neq 0$, which implies

$\varepsilon_1 = \alpha + \mu + \beta I_*, \varepsilon_2 = \gamma + \mu, \varepsilon_3 = \delta + \mu$ and $\varepsilon_4 = \mu + \sigma\beta I_*$. Thus,

$$J_* = \begin{bmatrix} -\varepsilon_1 & 0 & -\beta S_* & 0 & 0 \\ \beta I_* & -\varepsilon_2 & \beta(S_* + \sigma V_*) & 0 & \sigma\beta I_* \\ 0 & \gamma & -\varepsilon_3 & 0 & 0 \\ 0 & 0 & \delta & -\mu & 0 \\ \alpha & 0 & -\sigma\beta V_* & 0 & -\varepsilon_4 \end{bmatrix}. \quad \text{(S59)}$$

Its characteristic polynomial is

$$|\lambda I - J_*| = (\lambda + \mu)\{(\lambda + \varepsilon_4)[(\lambda + \varepsilon_1)(\lambda + \varepsilon_2)(\lambda + \varepsilon_3) + \gamma\beta[\beta I_* S_* - (\lambda + \varepsilon_1)((S_* + \sigma V_*)]$$

$$+\gamma\beta^2[(\lambda + \varepsilon_1)\sigma^2 I_* V_* + \alpha\sigma I_* S_*]\}\ , \quad \text{(S60)}$$

Or

$$|\lambda I - J_*| = (\lambda + \mu)\{\lambda^4 + (\varepsilon_1 + \varepsilon_2 + \varepsilon_3 + \varepsilon_4)\lambda^3 + [\varepsilon_4(\varepsilon_1 + \varepsilon_2 + \varepsilon_3) + \varepsilon_1(\varepsilon_2 + \varepsilon_3) + \varepsilon_2\varepsilon_3$$

$$-\gamma\beta(S_* + \sigma V_*)]\lambda^2 + [\varepsilon_2\varepsilon_3\varepsilon_4 + \varepsilon_1(\varepsilon_2\varepsilon_3 + \varepsilon_2\varepsilon_4 + \varepsilon_3\varepsilon_4) - \gamma\beta(\varepsilon_1 + \varepsilon_4)(S_* + \sigma V_*) + \gamma\beta^2 S_* I_* +$$

$$\gamma\beta^2\sigma^2 V_*]\lambda + \varepsilon_1\varepsilon_2\varepsilon_3\varepsilon_4 - \gamma\beta(S_* + \sigma V_*)\varepsilon_1\varepsilon_4 + \gamma\sigma^2\beta^2\varepsilon_1 I_* V_* + \gamma\beta^2\varepsilon_4 I_* S_* + \alpha\gamma\sigma\beta^2 I_* S_*\}. \quad \text{(S61)}$$

Using equations (S45) and (S59), we obtain

$$\gamma\beta(S_* + \sigma V_*) = \varepsilon_2\varepsilon_3 R_0^{end}, \quad \text{(S62)}$$

where

$$R_0^{end} = \frac{\gamma\beta\Lambda}{(\gamma+\mu)(\delta+\mu)(\alpha+\mu+\beta I_*)}[1 + \frac{\alpha\sigma}{(\mu+\sigma\beta I_*)}] \ . \tag{S63}$$

Again, substituting equation (S62) into equation (S60) yields

$$|\lambda I - J_*| = (\lambda + \mu)(\lambda + \varepsilon_4)\{(\lambda + \varepsilon_1)[(\lambda + \varepsilon_2)(\lambda + \varepsilon_3) - \varepsilon_2\varepsilon_3 R_0^{end} + \gamma\beta^2\sigma^2 I_* V_*] + $$

$$+\gamma\beta^2(1 + \alpha\sigma)I_* S_* \} \ . \tag{S64}$$

It follows from equation (S64) that the positive solution to characteristic equation (S64) comes from the equation

$$(\lambda + \varepsilon_1)[(\lambda + \varepsilon_2)(\lambda + \varepsilon_3) - \varepsilon_2\varepsilon_3 R_0^{end} + \gamma\beta^2\sigma^2 I_* V_*] + \gamma\beta^2(1 + \alpha\sigma)I_* S_* = 0 \tag{S65}$$

Considering that when dynamic system reaches stationary status, number of new cases $I_*$ is small. Thus, equation (S65) can be reduced to

$$(\lambda + \varepsilon_1)[(\lambda + \varepsilon_2)(\lambda + \varepsilon_3) - \varepsilon_2\varepsilon_3 R_0^{end}] = 0. \tag{S66}$$

Equation (S66) can be further reduced to

$$\lambda^3 + a_1\lambda^2 + a_2\lambda + a_3 = 0, \tag{S67}$$

where

$$a_1 = \varepsilon_1 + \varepsilon_2 + \varepsilon_3, a_2 = \varepsilon_1\varepsilon_2 + \varepsilon_1\varepsilon_3 + \varepsilon_2\varepsilon_3 - \varepsilon_2\varepsilon_3 R_0^{end}, a_3 = \varepsilon_1\varepsilon_2\varepsilon_3\left(1 - R_0^{end}\right).$$

Using Routh-Hurwitz stability criterion, we obtain that all roots in characteristic equation (S67) have negative real parts if and only if

$$H_1 = a_1 > 0, H_2 = \begin{vmatrix} a_1 & a_3 \\ 1 & a_2 \end{vmatrix} = a_1 a_2 - a_3 > 0, H_3 = \begin{vmatrix} a_1 & a_3 & 0 \\ 1 & a_2 & 0 \\ 0 & a_1 & a_3 \end{vmatrix} a_3(a_1 a_2 - a_3) > 0 \ . \tag{S68}$$

After some algebra, we can obtain from equation (S67) that

$$H_1 = \varepsilon_1 + \varepsilon_2 + \varepsilon_3 \ , \tag{S69}$$

$$H_2 = (\varepsilon_1 + \varepsilon_2 + \varepsilon_3)(\varepsilon_1\varepsilon_2 + \varepsilon_1\varepsilon_3) + (\varepsilon_2 + \varepsilon_3)\varepsilon_2\varepsilon_3\left(1 - R_0^{end}\right) , \tag{S70}$$

$$H_3 = \varepsilon_1\varepsilon_2\varepsilon_3\left(1 - R_0^{end}\right)H_2 \ . \tag{S71}$$

Recall that

$$\varepsilon_1 = \alpha + \mu + \beta I_*, \varepsilon_2 = \gamma + \mu, \varepsilon_3 = \delta + \mu \ ,$$

which implies

$$H_1 > 0 \ \text{ for all cases.} \tag{S72}$$

It is clear that if $R_0^{end} < 1$ then

$$H_2 > 0 \ , H_3 > 0 \ .$$

Using Routh-Hurwitz stability criterion, we obtain that all roots in characteristic equation (S67) have negative real parts. Therefore, if $R_0^{end} < 1$ then the endemic equilibrium point is stable.

Next we consider $R_0^{end} = 1$. In this case, we have

$a_3 = 0$ , which implies that

$H_3 = 0$ and

Routh-Hurwitz stability criterion is violated. Thus, the endemic equilibrium point is unstable.

Finally, we consider $R_0^{end} > 1$. The condition

$$R_0^{end} > 1$$

Implies that

$$a_3 = \varepsilon_1\varepsilon_2\varepsilon_3\left(1 - R_0^{end}\right) < 0 \ . \tag{S73}$$

Since

$H_3 = \varepsilon_1\varepsilon_2\varepsilon_3\left(1 - R_0^{end}\right)H_2$ , which implies

that $H_2$ and $H_3$ have opposite sign. Therefore, if $R_0^{end} > 1$ then again Routh-Hurwitz stability criterion is violated and the endemic equilibrium point is unstable

Therefore, in summary, the **endemic equilibrium point can be classified as three cases:**

(1) when $R_0^{end} < 1$, the endemic equilibrium point is classified as a asymptotically stable node;

(2) when $R_0^{end} = 1$, the endemic equilibrium point is classified as an unstable node; and

(3) when $R_0^{end} > 1$, the endemic equilibrium point is classified as an unstable saddle point.

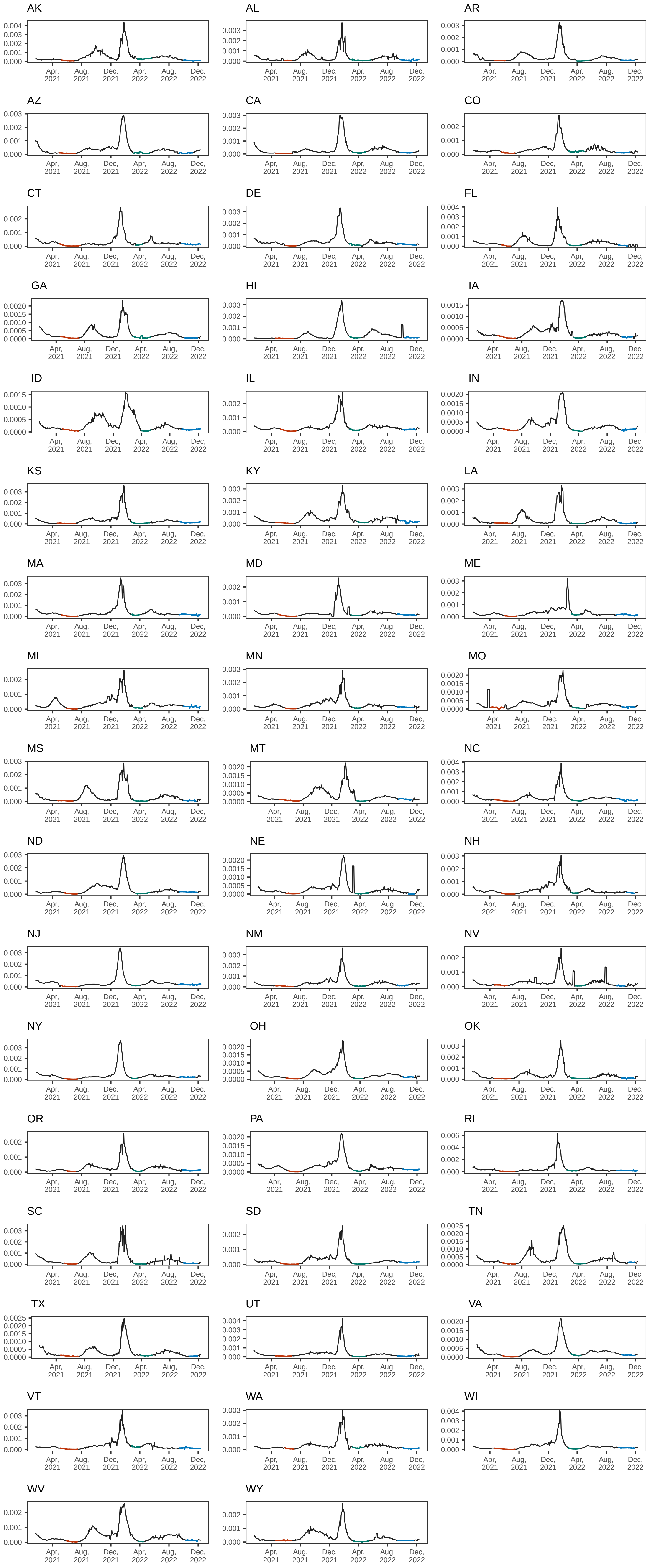

AK
AL
AR
AZ
CA
CO
CT
DE
FL
GA
HI
IA
ID
IL
IN
KS
KY
LA
MA
MD
ME
MI
MN
MO
MS
MT
NC
ND
NE
NH
NJ
NM
NV
NY
OH
OK
OR
PA
RI
SC
SD
TN
TX
UT
VA
VT
WA
WI
WV
WY
Apr, 2021
Aug, 2021
Dec, 2021
Apr, 2022
Aug, 2022
Dec, 2022

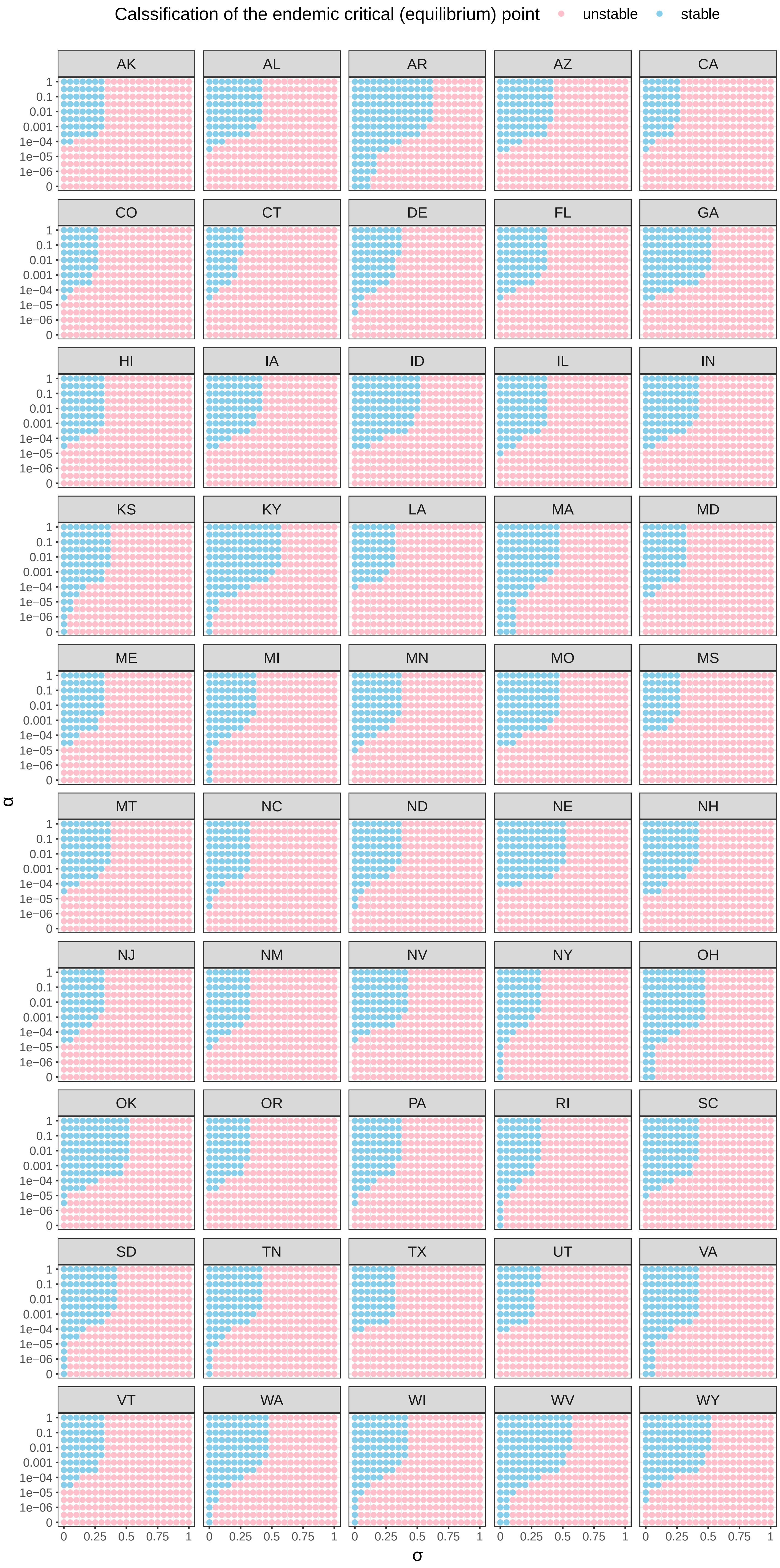
Calssification of the endemic critical (equilibrium) point
unstable
stable
AK
AL
AR
AZ
CA
CO
CT
DE
FL
GA
HI
IA
ID
IL
IN
KS
KY
LA
MA
MD
ME
MI
MN
MO
MS
MT
NC
ND
NE
NH
NJ
NM
NV
NY
OH
OK
OR
PA
RI
SC
SD
TN
TX
UT
VA
VT
WA
WI
WV
WY
1
0.1
0.01
0.001
1e−04
1e−05
1e−06
0
α
0
0.25
0.5
0.75
1
σ

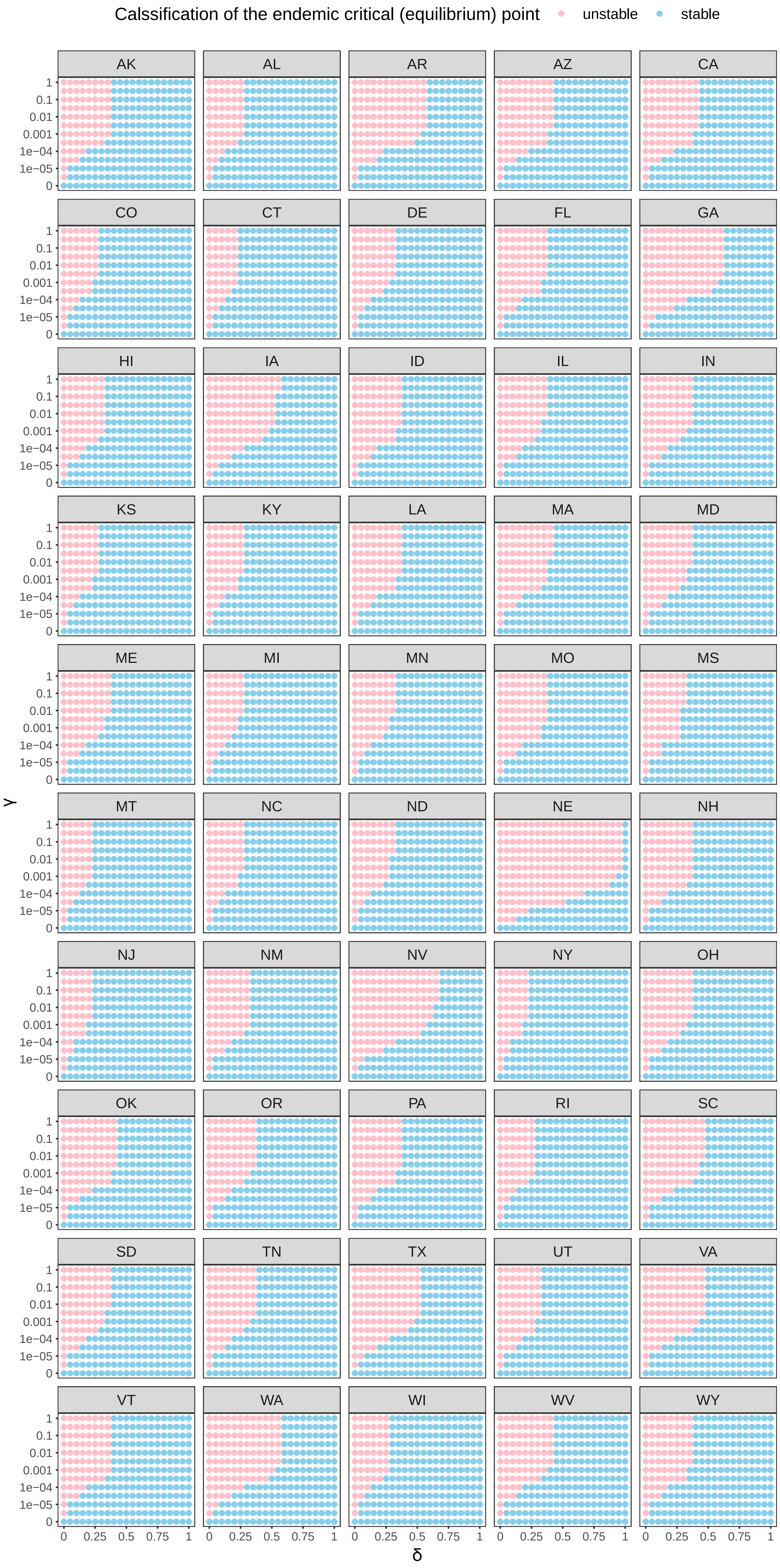
Calssification of the endemic critical (equilibrium) point
unstable
stable
AK
AL
AR
AZ
CA
CO
CT
DE
FL
GA
HI
IA
ID
IL
IN
KS
KY
LA
MA
MD
ME
MI
MN
MO
MS
MT
NC
ND
NE
NH
NJ
NM
NV
NY
OH
OK
OR
PA
RI
SC
SD
TN
TX
UT
VA
VT
WA
WI
WV
WY
γ
1
0.1
0.01
0.001
1e−04
1e−05
0
δ
0
0.25
0.5
0.75
1

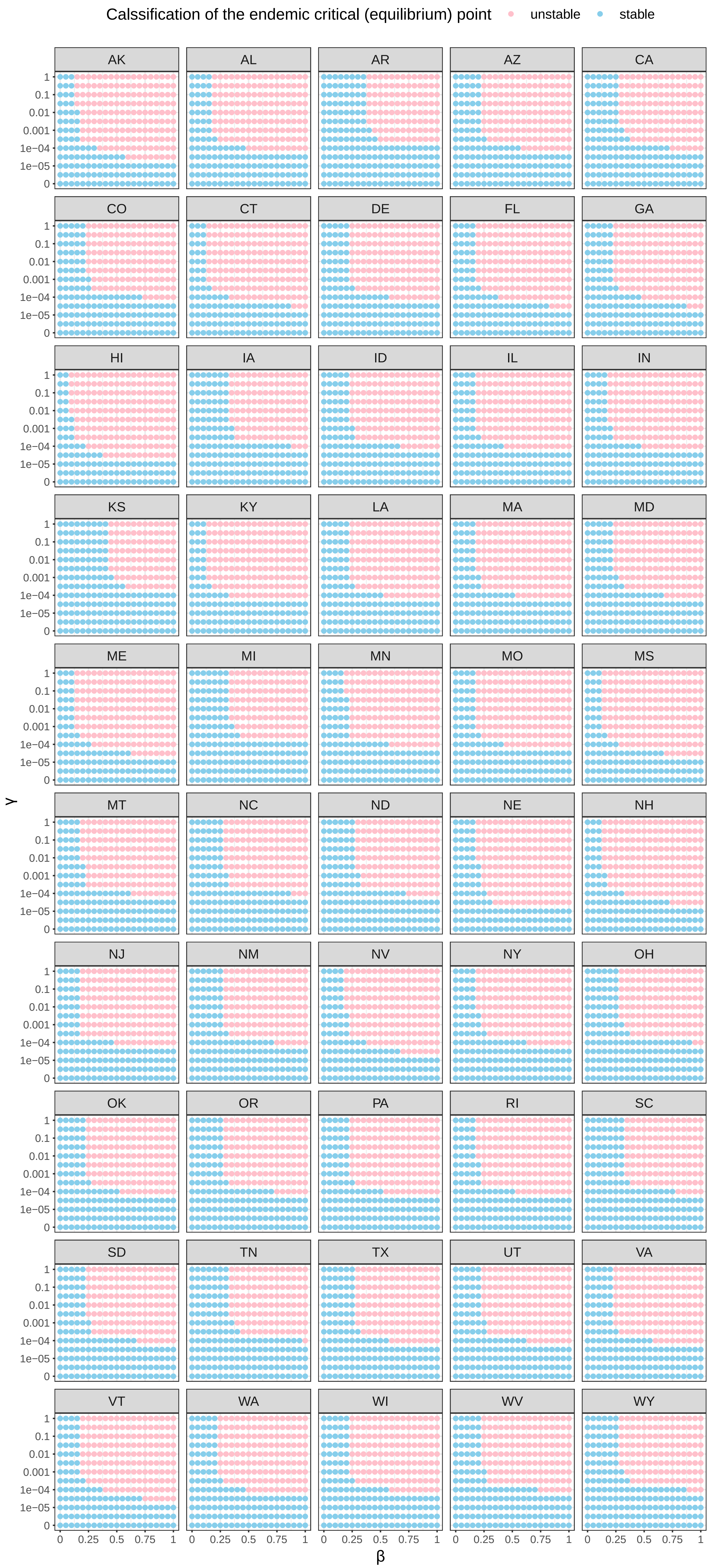

Calssification of the endemic critical (equilibrium) point
unstable
stable
AK
AL
AR
AZ
CA
CO
CT
DE
FL
GA
HI
IA
ID
IL
IN
KS
KY
LA
MA
MD
ME
MI
MN
MO
MS
MT
NC
ND
NE
NH
NJ
NM
NV
NY
OH
OK
OR
PA
RI
SC
SD
TN
TX
UT
VA
VT
WA
WI
WV
WY
γ
1
0.1
0.01
0.001
1e−04
1e−05
0
β
0
0.25
0.5
0.75
1

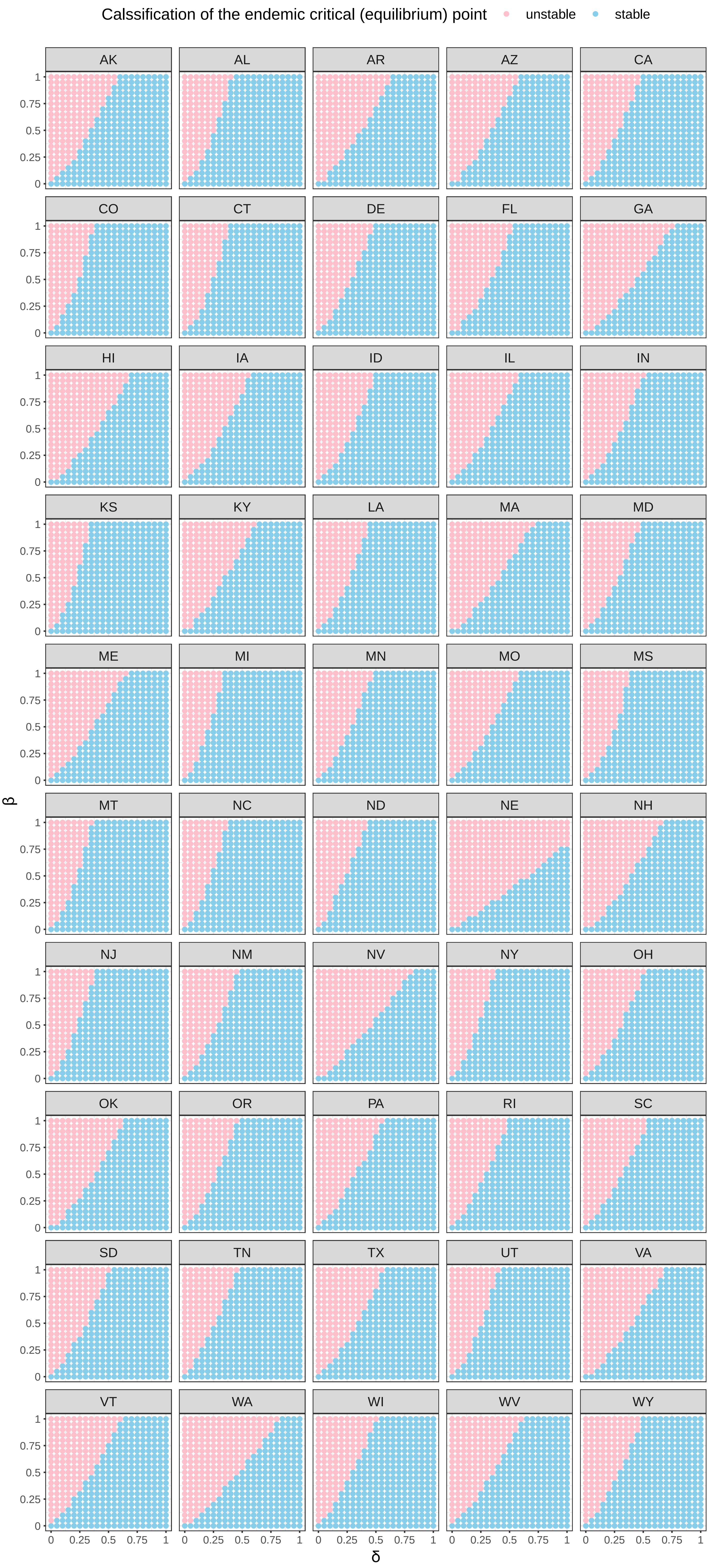
Calssification of the endemic critical (equilibrium) point
unstable
stable
AK
AL
AR
AZ
CA
CO
CT
DE
FL
GA
HI
IA
ID
IL
IN
KS
KY
LA
MA
MD
ME
MI
MN
MO
MS
MT
NC
ND
NE
NH
NJ
NM
NV
NY
OH
OK
OR
PA
RI
SC
SD
TN
TX
UT
VA
VT
WA
WI
WV
WY
β
δ
0
0.25
0.5
0.75
1

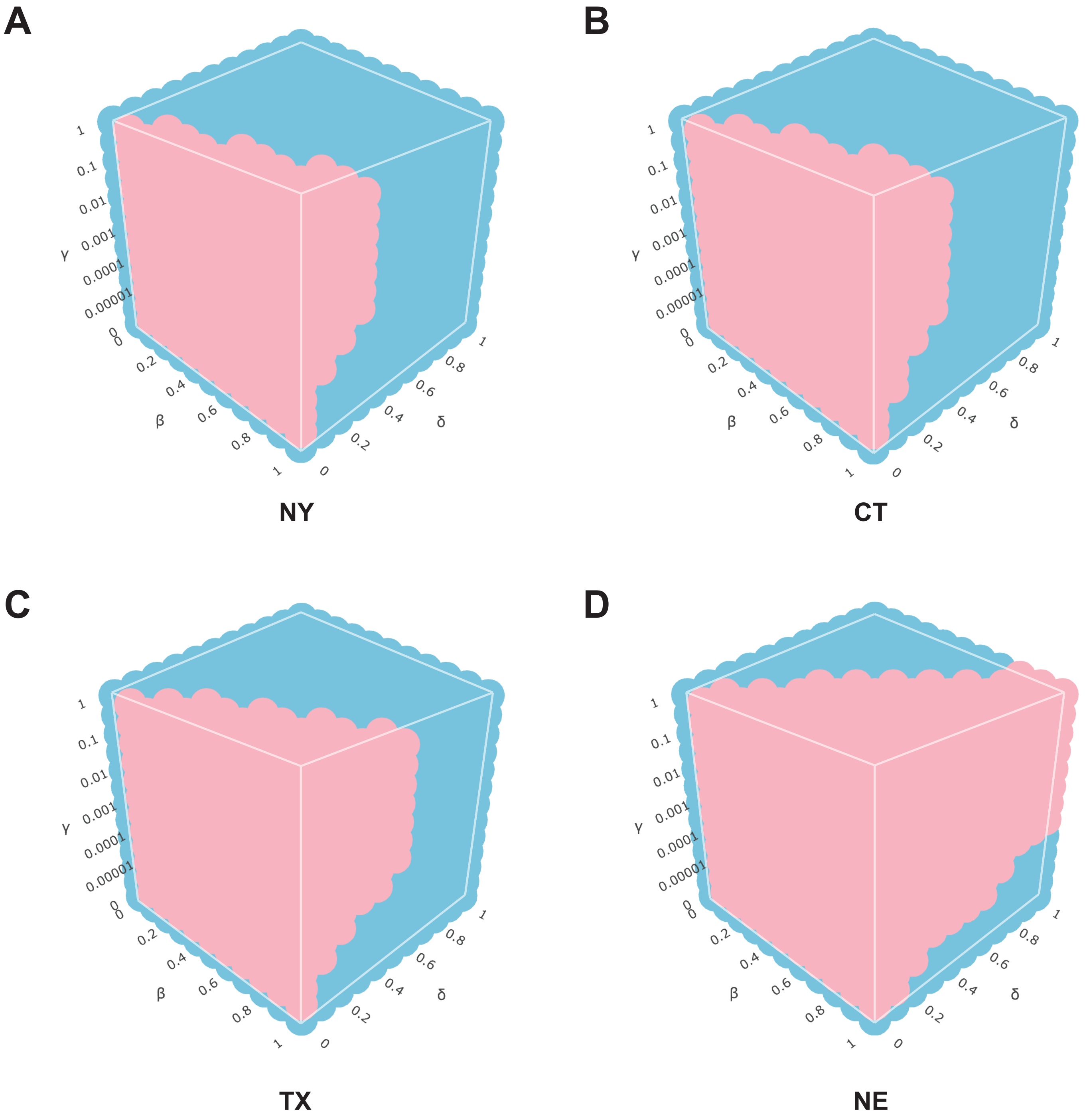
A
B
C
D
NY
CT
TX
NE
γ
β
δ
1
0.1
0.01
0.001
0.0001
0.00001
0
0.2
0.4
0.6
0.8
